# Single-cell transcriptomics identifies different immune signatures between macrophage activation-like syndrome and immune paralysis in sepsis

**DOI:** 10.1101/2023.03.17.23287390

**Authors:** Inge Grondman, Valerie A.C.M. Koeken, Athanasios Karageorgos, Wenchao Li, Nikolaos Antonakos, Bowen Zhang, Georgia Damoraki, Cheng-Jian Xu, Evangelos J. Giamarellos-Bourboulis, Yang Li, Mihai G. Netea

**Affiliations:** Department of Internal Medicine and Radboud Center for Infectious Diseases, Radboud University Medical Center, Nijmegen, the Netherlands; Department of Computational Biology for Individualised Medicine, Centre for Individualised Infection Medicine (CiiM), a joint venture between the Hannover Medical School and the Helmholtz Centre for Infection Research, Hannover, Germany; TWINCORE, Centre for Experimental and Clinical Infection Research, a joint venture between the Hannover Medical School and the Helmholtz Centre for Infection Research, Hannover, Germany; 4th Department of Internal Medicine, National and Kapodistrian University of Athens, Medical School, Greece; Hellenic Institute for the Study of Sepsis, Athens, Greece; Department of Immunology and Metabolism, Life and Medical Sciences Institute (LIMES), University of Bonn, Germany

## Abstract

Different immune phenotypes characterize sepsis patients, including hyperinflammation and/or immunosuppression, but the biological mechanisms driving this heterogeneity remain largely unknown. We used single-cell RNA sequencing to profile circulating leukocytes of healthy controls and sepsis patients classified as either *hyperinflammatory* (macrophage activation-like syndrome [MALS]), *immune paralysis*, or *unclassified* (when criteria for neither of these two immune subgroups were applicable). Pronounced differences were detected in the transcriptional signature of monocytes from sepsis patients, with clear distinction between MALS and immune paralysis patients. Unsupervised clustering analysis revealed the existence of MALS-specific monocyte clusters, as well as one sepsis-specific monocyte cluster that was linked to disease severity. In separate cohorts, urosepsis was characterized by heterogeneous MALS and immunosuppression monocyte signatures, while MALS-specific monocyte clusters showed overlapping transcriptional signatures with severe COVID-19. In conclusion, our findings shed light on the heterogeneous immune landscape underlying sepsis, and provide opportunities for patient stratification for future therapeutic development.

## Introduction

Sepsis is a life-threatening disease in which a dysregulated host response (either exaggerated or deficient) against a severe infection is a crucial component of the pathogenesis (1–3). Factors including age, source of infection, comorbidities and disease course influence its outcome, adding additional complexity to patient stratification based on individual host responses (4). The heterogeneous nature of sepsis is considered to be a major barrier to improving treatment, and this may explain why earlier randomized-controlled trials of immunomodulating therapies in sepsis have failed to show clinical benefits (5–7).

Different immunological and clinical phenotypes can be recognized in sepsis patients. Some patients present with an overwhelming hyperinflammatory state, more often observed during early stages of disease. These patients show clinical symptoms and features similar to macrophage activation syndrome (MAS), a form of secondary hemophagocytic lymphohistocytosis (sHLH), including signs of hepatobiliary dysfunction and disseminated intravascular coagulation. Although bone marrow hemophagocytosis is one of the original criteria in the diagnosis of primary (genetic) HLH, the diagnosis of secondary MAS may be done using criteria other than traits of hemophagocytosis in bone marrow aspirates; this is why this state is preferably called macrophage activation-like syndrome (MALS) to describe a hyperinflammatory subgroup of sepsis patients (8, 9). MALS is associated with high mortality, and an increased circulating concentration of ferritin above 4,420 ng/mL has been proposed as an important biomarker for this immune phenotype (8). On the other hand, patients may develop a profound state of immunosuppression or so-called sepsis-induced immune paralysis, characterized by low monocytic HLA-DR expression on monocytes and clinically recognized by their increased vulnerability to opportunistic infections (10–12).

The identification of the subgroups of patients with distinct signatures of immune dysregulation is a promising approach for patient stratification-based trials and personalized treatment strategies in sepsis. A recent study has shown that high ferritin levels and decreased monocytic HLA-DR expression can be used to characterize patients with distinct features of MALS or immune paralysis (13). However, the underlying pathophysiological mechanisms driving these entities, so-called endotypes, are still largely unknown. Our goal was to provide new insights in the cellular and molecular basis of immune dysregulation of sepsis driving these endotypes, as it may reveal more specific disease signatures. This has the potential to further facilitate personalized care and pre-enrichment strategies in sepsis patients, as well as the choice for the immunotherapeutic approach most likely to be successful. Therefore, in this study we profiled the transcriptome of circulating immune cells on a single-cell level in selected subgroups of sepsis patients with different immunological subclasses, namely those with extreme hyperinflammatory (MALS) and immunosuppressive states.

## Results

### Sample overview and cell proportion changes in sepsis immune endotypes

Peripheral blood mononuclear cells (PBMCs) were isolated from sepsis patients and healthy controls, after which single-cell RNA-sequencing (scRNA-seq) was performed to characterize the immune signature of sepsis and specific immunological subgroups. Sepsis patients admitted to one of the recruiting hospitals in Greece were screened for immunological dysfunction and classified into the *macrophage activation-like syndrome* (MALS) immune endotype if circulating ferritin concentrations were >4,420 ng/mL; *immune paralysis* in case HLA-DR expression was below 5,000 monoclonal-antibodies binding sites per monocyte (Quantibrite methodology) and ferritin concentration was ≤4,420 ng/mL; and as *unclassified* if neither MALS or immune paralysis criteria were applicable. One sample was identified as an outlier based on the uniform manifold approximation and projection (UMAP) plot (**Supplemental Figure 1A**), and therefore excluded from further analysis, leaving us with biological samples of 16 sepsis (5 MALS, 6 unclassified, and 5 immune paralysis) and 6 healthy individuals for further analysis (**Figure 1A**).

**Figure 1:**
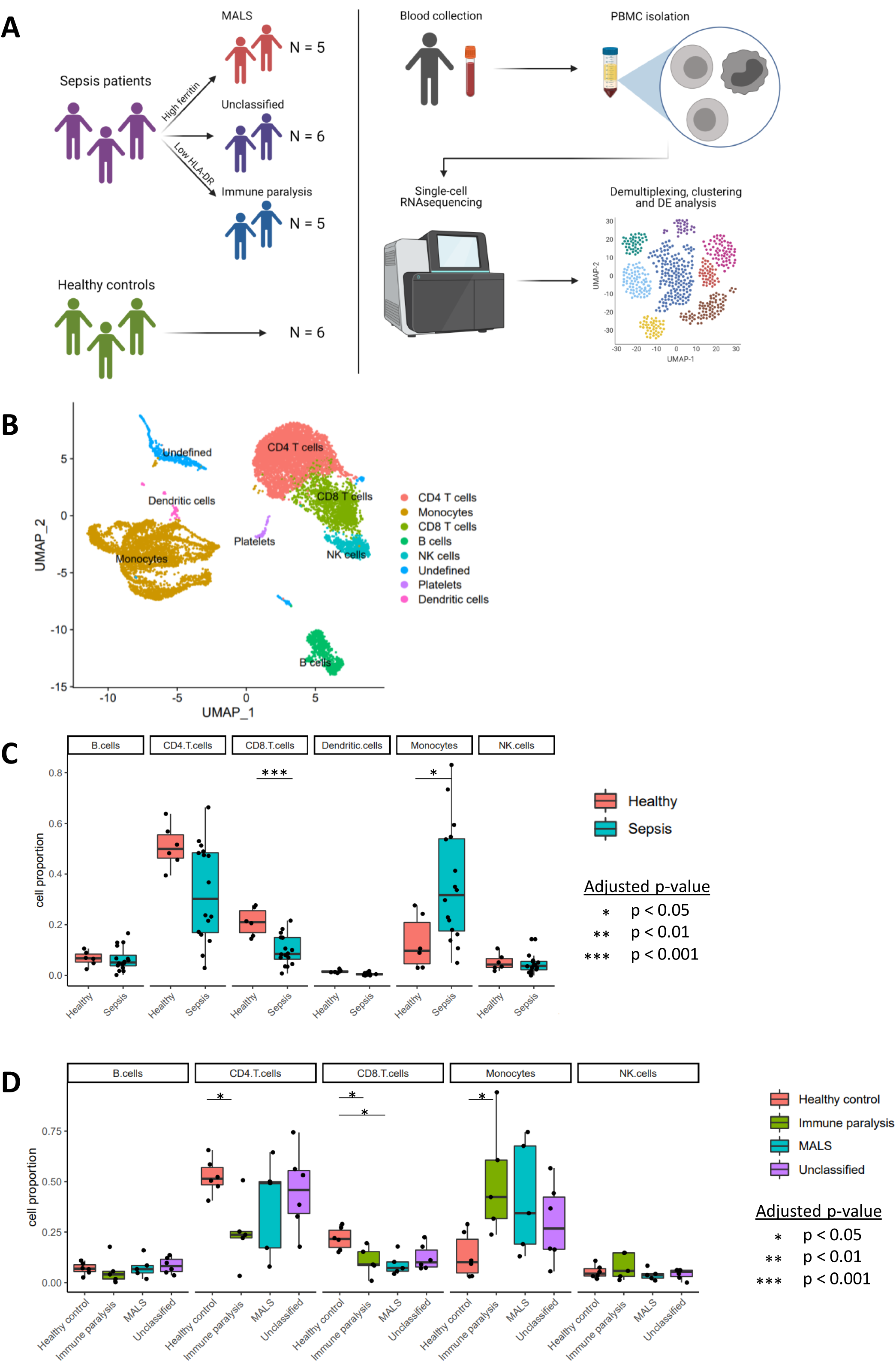
Overview of the study approach and single-cell RNA-sequencing data. (**A**) General overview of the study. Peripheral blood mononuclear cells were isolated from EDTA blood from 16 sepsis patients (5 MALS, 6 unclassified and 5 immune paralysis) and 6 healthy controls, after which single-cell RNA-sequencing was performed using 10X Genomics technology. Created with BioRender.com. (**B**) Uniform manifold approximation and projection (UMAP) plot of 13,059 peripheral blood mononuclear cells from 22 samples (6 healthy controls and 16 sepsis patients). The clusters are annotated based on marker gene expression. (**C-D**) Boxplots of cell proportions for the main cell types, comparing healthy controls to sepsis patients (**C**) or comparing healthy controls to immune paralysis, MALS and unclassified patients (**D**). False discovery rate (FDR) adjusted p-value<0.05 was considered to be statistically significant.

The majority of sepsis patients included in this study suffered from septic shock (14/16, 88%). The main cause of sepsis was pneumonia (13/16, 81%), either community-acquired, hospital-acquired, or associated with mechanical ventilation, based on clinical predefined criteria. Cultures remained sterile for more than half of the patients (10/16, 63%). Five patients (32%) had positive cultures with bacteria. In one patient, viral cause of sepsis was identified (Human coronavirus OC43). Overall, no substantial differences in patient characteristics were observed between the immunological subgroups, except for a significant higher Sequential Organ Failure Assessment (SOFA) score in patients with MALS, compared to patients with immune paralysis and those defined as unclassified. Baseline and clinical characteristics of all sepsis patients and immunological subgroups are summarized in **Table 1**. In addition, this study included a group of healthy individuals (n=6; 4 females, 2 males) with a slightly lower age (median of 52 years [IQR44-56] in the control group vs. 66 years [IQR 59-85] in the sepsis group, p-value 0.013), but similar BMI (healthy controls median 27 [IQR 24-32] vs. 27 [IQR 22-29]) compared to the total sepsis population.

**Table 1.** Baseline and clinical characteristics of sepsis patients and the immunological subgroups

|  | **Total**<br>**(n=16)** | **MALS**<br>**(n=5)** | **Immune paralysis**<br>**(n=5)** | **Unclassified**<br>**(n=6)** | **P-**<br>**value\*** |
| --- | --- | --- | --- | --- | --- |
| Age, years | 66 (59-85) | 65 (62-80) | 65 (45-91) | 72 (45-81) | NS |
| Gender (n,%) |  |  |  |  |  |
| Male | 12 (75) | 4 (80) | 3 (60) | 5 (83) | NS |
| Female | 4 (25) | 1 (20) | 2 (40) | 1 (17) | NS |
| Body Mass Index, kg/m<sup>2</sup> | 27.1<br>(21.6-29.4) | 29.4<br>(22.0-35.3) | 21.5<br>(21.0-25.9) | 27.2<br>(25.4-29.4) | NS |
| Comorbidities (n,%) |  |  |  |  |  |
| Diabetes mellitus | 6 (38) | 2 (40) | 2 (40) | 2 (33) | NS |
| Heart failure | 5 (31) | 1 (20) | 1 (20) | 3 (50) | NS |
| Coronary heart disease | 7 (44) | 3 (60) | 0 (0) | 4 (67) | NS |
| Chronic renal disease | 1 (6) | 0 (0) | 0 (0) | 1 (17) | NS |
| Chronic obstructive pulmonary disease | 5 (31) | 1 (20) | 1 (20) | 3 (50) | NS |
| History of hematological/solid malignancies | 2 (13) | 1 (20) | 1 (20) | 0 (0) | NS |
| Charlson comorbidity index (CCI) | 5 (3-8) | 5 (4-6) | 6 (1-7) | 7 (3-8) | NS |
| Source of infection (n,%) |  |  |  |  |  |
| Acute cholangitis | 1 (6) | 1 (20) | 0 (0) | 0 (0) | NS |
| Primary bacteremia | 2 (13) | 1 (20) | 1 (20) | 0 (0) | NS |
| Community acquired pneumonia | 6 (38) | 0 (0) | 3 (60) | 3 (50) | NS |
| Health care associated pneumonia | 4 (25) | 3 (60) | 0 (0) | 1 (17) | NS |
| Ventilator associated pneumonia | 3 (19) | 0 (0) | 1 (20) | 2 (33) | NS |
| Septic shock (n,%) | 14 (88) | 5 (100) | 4 (80) | 5 (83) | NS |
| SOFA score | 12 (8-15) | 15 (15-17) | 8 (6-14) | 11 (8-12) | **0.009<sup>#</sup>** |
| Mortality 28 days (n,%) | 4 (25) | 3 (60) | 0 (0) | 1 (17) | NS |
| Data are presented as median with interquartile range (IQR) or n (%).<br>* The difference between the 3 subgroups (MALS, immune paralysis and unclassified) was statistically tested with the Kruskal-Wallis test for continuous data, or with the Fisher-Freeman-Halton test for nominal data. A P-value of <0.05 is considered statistical significant.<br># P-value adjusted by the Bonferroni correction for multiple testing: 0.023 for immune paralysis vs. MALS and 0.021 for unclassified vs. MALS<br>Abbreviations: MALS; macrophage activation-like syndrome, CCI; Charlson comorbidity index, APACHE; acute physiology and chronic health evaluation, NS; not significant, SOFA; sequential organ failure assessment, IQR; interquartile range. |  |  |  |  |  |

PBMCs were sequenced using 10X Genomics technology in batches of 5 samples per experiment, and no major batch effects were observed in these data. After filtering for singlets, number of features per cell and percentage of mitochondrial reads, more than 13,000 cells across 22 samples from individual participants were left for further analyses. Using UMAP clustering in combination with marker gene expression, all major cell types expected in the mononuclear compartment of the blood were identified (**Figure 1B** and **Supplemental Figure 1B**). Cell subset analysis revealed a shift in circulating immune cell proportions in sepsis, which was characterized by a significant increase in monocytes (Dirichlet regression analysis, false discovery rate [FDR] adjusted p-value=0.04) and a decrease in CD8 T cells (adjusted p-value=5.5×10^-4^, **Figure 1C**). This shift was more pronounced in the immune paralysis group, in whom a significant decrease in CD4 and CD8 T cell proportions was observed (adjusted p-value=0.04 and adjusted p-value=0.02, respectively), as well as an increase in the monocyte fraction (adjusted p-value=0.02, **Figure 1D**). These changes in cell proportions in different immune endotypes underline the dysregulation of the host response in sepsis.

### Cell-type specific transcriptomic changes in sepsis

To define if the dysregulated immune response in sepsis is caused solely by differences in cell proportions or also through changes in transcriptional programs, we compared the gene expression signatures per cell type between sepsis patients and healthy controls. Of the six main cell types studied here (monocytes, dendritic cells, NK cells, CD4 T cells, CD8 T cells and B cells; **Supplemental Figure 2**), differences in transcriptome between healthy individuals and sepsis patients were most noticeable in monocytes (**Figure 2A**) and CD4 T cells (**Figure 2B**). An overview of the number of differentially expressed genes in various cell types is shown in **Figure 2C**. In the CD4 T cells, 29% of the genes included in the analysis were significantly altered in sepsis (adjusted p-value<0.05), while in the monocytes, 12% of the transcriptome was differentially regulated upon disease. Among the most upregulated genes in the CD4 T cells of sepsis patients were *SOCS3*, which is a suppressor of cytokine signaling, and *FOS* and *JUN*, which form a transcription factor regulating cell proliferation and differentiation. In monocytes, the most upregulated gene in sepsis was *THBS1*, which is a glycoprotein involved in platelet aggregation and angiogenesis, while the most downregulated genes are *HLA-DQA1* and *HLA-DQB1*, which are both HLA class II molecules used for antigen presentation. Further enrichment using the KEGG pathway database revealed that genes upregulated in sepsis were enriched in pathways involved in oxidative phosphorylation (in CD4 T cells and monocytes) and TNF signaling (in CD8 T cells, monocytes and NK cells; **Figure 2D**), and these pathways were also enriched in a sensitivity analysis corrected for age and sex (**Supplemental Figure 3**). The number of pathways enriched based on the downregulated genes in sepsis was much lower, but included antigen processing and presentation (in B cells and monocytes; **Figure 2E**), which was also observed in a sensitivity analysis corrected for age and sex (**Supplemental Figure 3**). GO terms enrichment analysis revealed NADH dehydrogenase activity to be enriched in CD4 T cells based on the upregulated genes, and again pathways involved in antigen presentation (MHC protein complex binding) were downregulated in B cells and monocytes (**Supplemental Figure 4**). Since we observed a high number of differentially expressed genes, both up as well as downregulated, in the monocytes from sepsis patients, we built up a monocyte-centric ligand-receptor interaction atlas between the main cell types. This analysis revealed that monocytes in sepsis interact with a variety of other cell types, showing the highest number of interactions with other monocytes (**Figure 2F**). We also observed a crosstalk pathway, in which the decoy receptor IL-1 receptor type II (*IL1R2*) on monocytes potentially interacted and neutralized IL-1β (*IL1B*) from dendritic cells, which is most likely relevant for sepsis. Together, these data identify altered gene expression profiles in sepsis, especially in CD4 T cells and monocytes.

**Figure 2:**
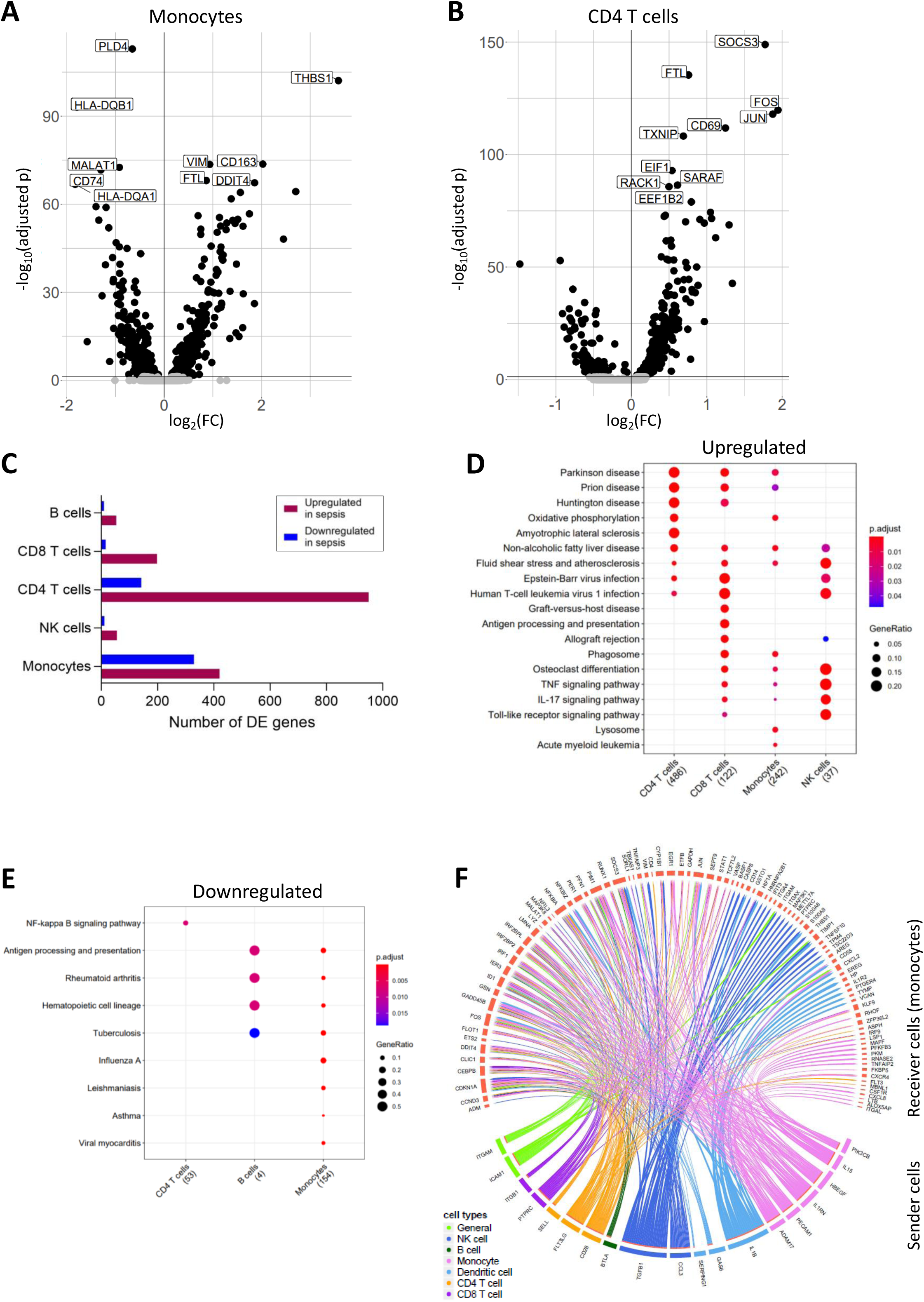
Sepsis-specific immune signatures compared to healthy controls. Volcano plots showing results from differential expression analysis (Wilcoxon rank-sum tests) between sepsis patients and healthy controls in the monocyte (**A**) and CD4 T cell subset (**B**). The -log_10_(Bonferroni adjusted p-value) is visualized on the y-axis (adjusted p-value<0.05 is considered to be statistically significant) and the log_2_(fold change) is visualized on the x-axis. (**C**) Number of differentially expressed genes per cell type with an adjusted p-value<0.05. Upregulated genes (higher in sepsis patients compared to healthy controls) are visualized in purple, downregulated genes in blue. (**D**-**E**) Enrichment analyses based on upregulated genes (**D**) or downregulated genes (**E**) using the KEGG database. (**F**) Monocyte-centric ligand-receptor interaction atlas using the NicheNet algorithm with monocyte as the receiver cell type and the other main cell types as sender cells.

### Sepsis patients with MALS or immune paralysis show profound transcriptional differences in monocytes

To understand the immunological basis which distinguishes MALS from immune paralysis patients, we compared gene expression in immune cells between these two patient groups. Excluding the dendritic cells from this analysis due to low cell numbers, the differentially expressed genes of the five remaining cell types were calculated (**Supplemental Figure 5**). Among these five cell types, the most pronounced differences were observed in the monocyte population (**Figure 3A**). Among the differentially expressed genes in monocytes between MALS and immune paralysis patients (**Figure 3B**), *FTL*, which is a subunit of ferritin, was increased in monocytes from MALS compared to immune paralysis patients (**Figure 3C**), confirming the high circulating ferritin concentrations used to define the immune class MALS. Further analysis based on the genes higher expressed in MALS patients revealed an enrichment in apoptosis (in CD4 T cells, B cells, monocytes and NK cells) and phagosome activation (in CD4 T cells, CD8 T cells, B cells and monocytes) pathways (KEGG pathways: **Figure 3D**). The GO terms enrichment analysis identified NADH hydrogenase activity to be enriched in monocytes from MALS patients (GO terms: **Supplemental Figure 6**). The number of pathways enriched based on immune paralysis-related genes was lower compared to the number of pathways related to MALS, and included mainly disease-related pathways e.g. HIV, primary immunodeficiency, T-cell leukemia and parathyroid hormone disease (KEGG pathways: **Figure 3E**).

**Figure 3:**
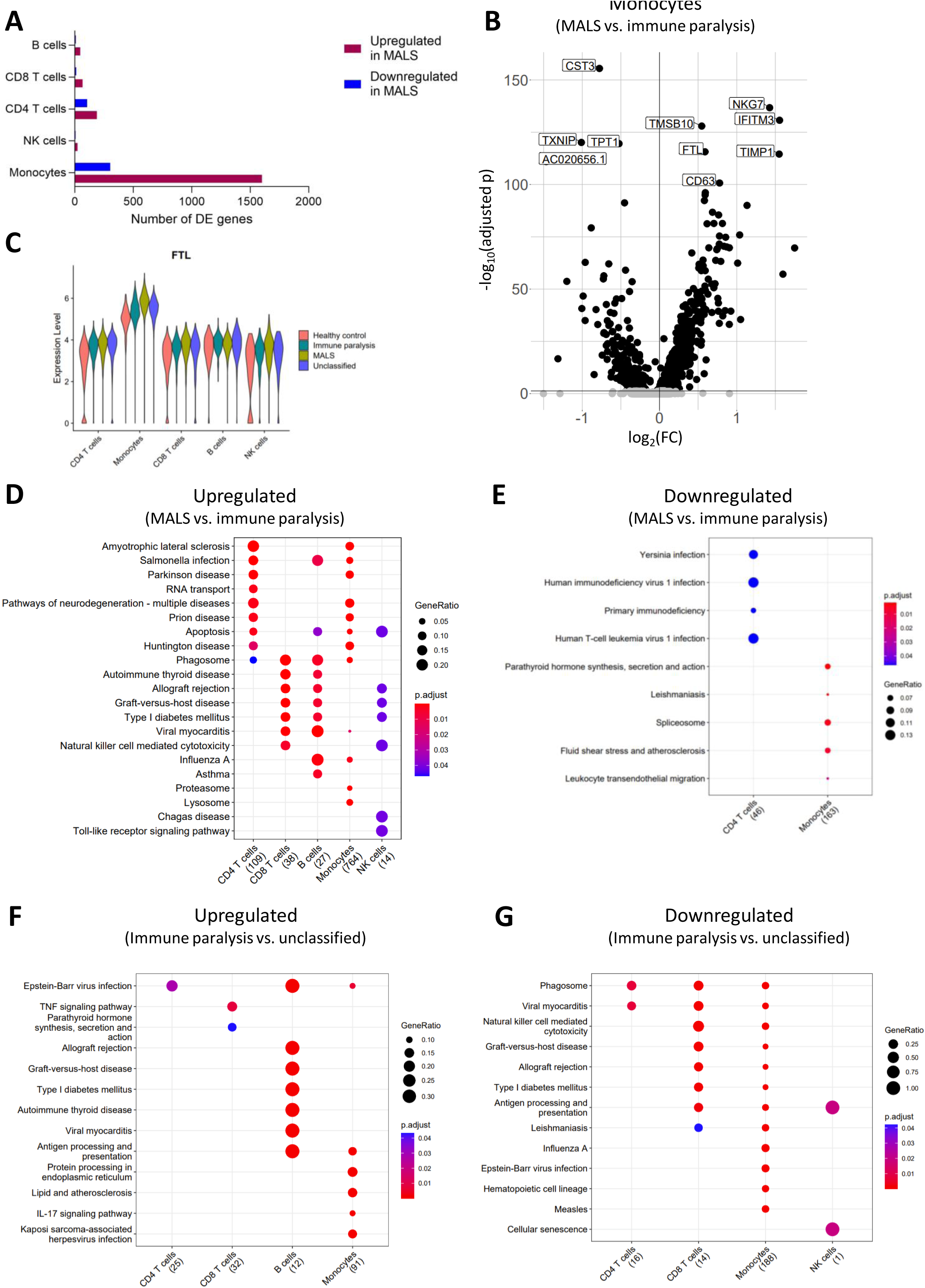
Differences in immune cell-states between MALS and immune paralysis patients. (**A**) Number of differentially expressed genes per cell type. Upregulated genes (higher in MALS patients compared to immune paralysis patients) are visualized in purple, downregulated genes in blue. (**B**) Volcano plot showing the differentially expressed genes (Wilcoxon rank-sum tests) between MALS and immune paralysis patients in monocytes. The -log_10_(Bonferroni adjusted p-value) is visualized on the y-axis (adjusted p-value<0.05 is considered to be statistically significant) and the log_2_(fold change) is visualized on the x-axis. (**C**) Violin plot of *FTL* expression per cell type separated by group. (**D**-**E**) Enrichment analyses based on upregulated genes (**D**) or downregulated genes (**E**) in MALS compared to immune paralysis patients using the KEGG database. (**F**-**G**) Enrichment analyses based on upregulated genes (**F**) or downregulated genes (**G**) in immune paralysis compared to unclassified patients using the KEGG database.

To characterize the immune paralysis endotype even further, we also compared immune paralysis patients to the unclassified group. Enrichment analysis based on differentially expressed genes identified pathways such as IL-17 signaling in monocytes based on upregulated genes and NK cell-mediated cytotoxicity in CD8 T cells and monocytes (**Figure 3F** and **3G**). Interestingly, antigen processing and presentation as well as MHC protein-related pathways were enriched based on both upregulated and downregulated genes, indicating active regulation of this pathway in immune paralysis (**Figure 3F**, **3G**, and **Supplemental Figure 7**).

Further, to understand the molecular drivers of the distinct sepsis immune endotypes, we performed a transcription factor (TF) enrichment analysis based on the differentially expressed genes comparing monocytes from one immune endotype to the other two groups. In total, 40 TFs were uniquely enriched in the MALS monocytes, 20 in monocytes from the unclassified immune endotype, and 30 in monocytes from immune paralysis patients (**Figure 4A**). The monocytes from unclassified patients were mainly driven by TFs related to interferon signaling, including *STAT1*, *STAT2*, *IRF2*, *IRF3*, *IRF5*, *IRF7* and *IRF9*. As unclassified patients most likely have the best outcome (13), this suggests that a proper activation of interferon pathways is a necessary component of host defense in sepsis. On the other hand, monocytes from immune paralysis patients are enriched in transcription factors from the *JUN* and *FOS* family, regulating monocyte differentiation (14), while MALS monocytes are enriched in transcription factors from the ETS family, including *ELF1*, *ELF2*, *ETV1* and *ETS2* (**Figure 4B**). The enrichment of these distinct transcription factor families shed light on the molecular regulation of monocytes in sepsis.

**Figure 4:**
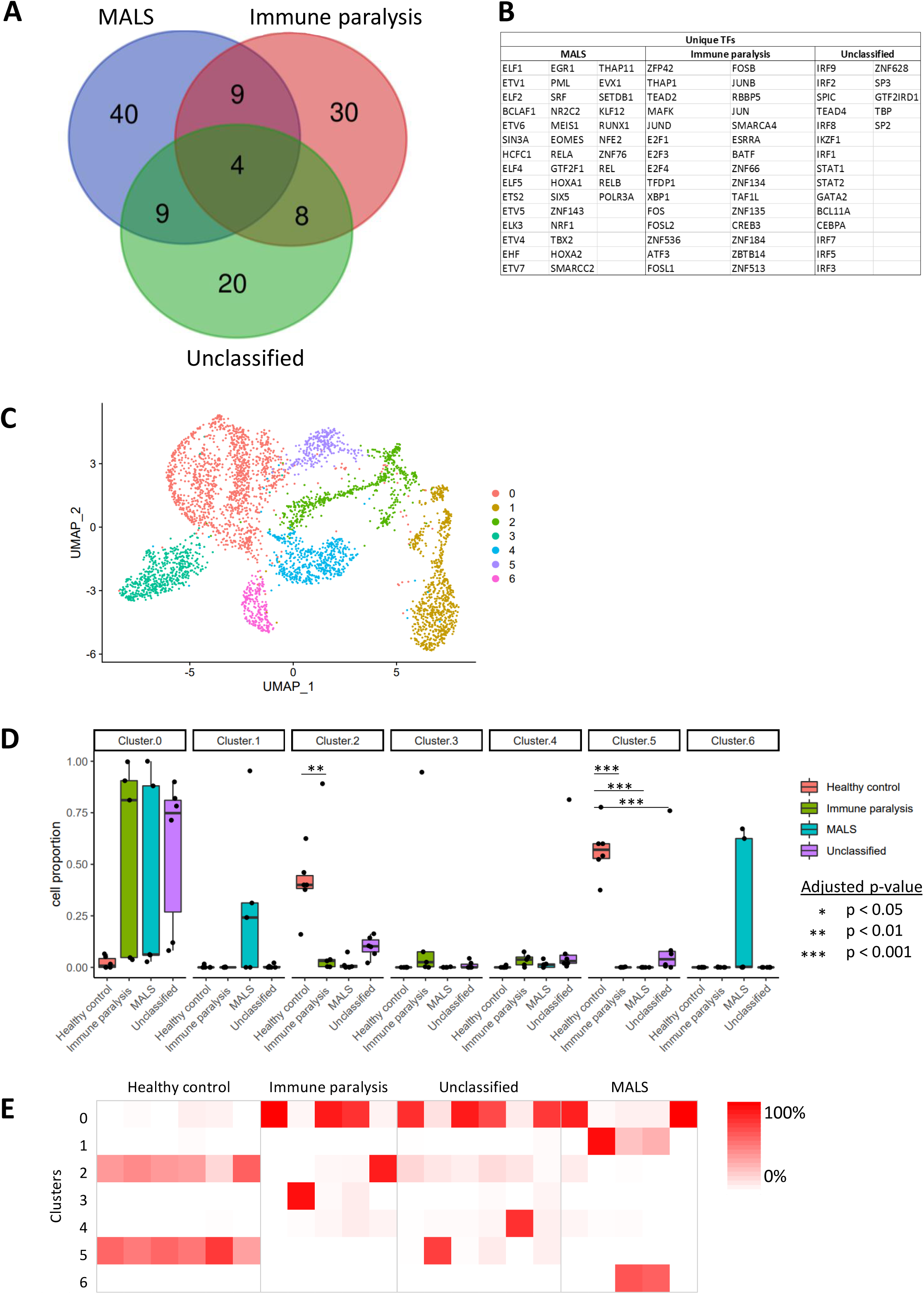
Monocyte signatures and enriched transcription factors in sepsis. (**A-B**) Enriched transcription factors based on differentially expressed genes between conditions in monocytes are visualized in a Venn diagram (**A**). The transcription factors that are uniquely enriched in only one condition are separately listed (**B**). (**C**) Uniform manifold approximation and projection (UMAP) plot of monocytes from 22 samples (6 healthy controls and 16 sepsis patients), identifying seven distinct monocyte clusters. (**D**) Boxplot of cell proportions of the seven monocyte clusters comparing healthy controls to immune paralysis, MALS and unclassified patients. FDR adjusted p-value<0.05 is considered to be statistically significant. (**E**) Visualization of monocyte cluster proportions among all 22 study participants, divided by endotype.

### Identification of monocyte subclusters related to sepsis and sepsis-specific immune endotypes

Considering the heterogeneity observed in the monocyte population, as well as the high number of differentially expressed genes, we decided to study this cell subset in more detail. UMAP clustering on the monocyte population revealed seven distinct subsets within this cell type (**Figure 4C**). These specific monocyte subsets were not equally distributed among the different sepsis immune endotypes (**Figure 4D**, **4E**, and **Supplemental Figure 8**). In this regard, monocytes of cluster 0 were present in large proportions in sepsis patients from all three immune endotypes, while monocytes from clusters 1 and 6 were identified only in MALS patients. In addition, monocytes from clusters 2 and 5 could be observed more frequently in healthy controls compared with sepsis patients (Dirichlet regression analysis, adjusted p-value<0.01). Clusters 3, 5 and 6 were predominantly made up by classical monocytes (high in *CD14* expression), while clusters 0, 1, 2 and 4 were made up of a mix of both classical and non-classical monocytes (high in *CD16* expression, **Supplemental Figure 8**). These analyses identify monocyte subsets that are unique to sepsis and to specific immune endotypes present in sepsis.

Since these monocyte subsets are present in different proportions across distinct sepsis immune endotypes, we aimed to understand the underlying differences between their transcriptional programs. Examining the marker genes specific for these monocyte clusters, we observed that several HLA genes were involved in separating the different monocytes populations, including *HLA-DRB5*, *HLA-DPB1*, *HLA-DQB1*, and *HLA-DMB* (**Figure 5A** and **Supplemental Figure 9**). Additional pathway analyses on the marker genes of each monocyte cluster revealed that monocytes from cluster 0, which are present in all sepsis immune endotypes, are characterized by genes related to IL-17 signaling and apoptosis (**Figure 5B**). In addition, monocytes from cluster 6, which could only be found in MALS patients, are characterized by genes associated to HIF-1 signaling and glycolysis. Other pathways enriched in specific monocyte clusters include antigen processing and presentation (cluster 0, 1, 2, and 5), phagosome (cluster 1, 2, 5, and 6), and hematopoietic cell lineage (cluster 1, 2, 4, 5, and 6).

**Figure 5:**
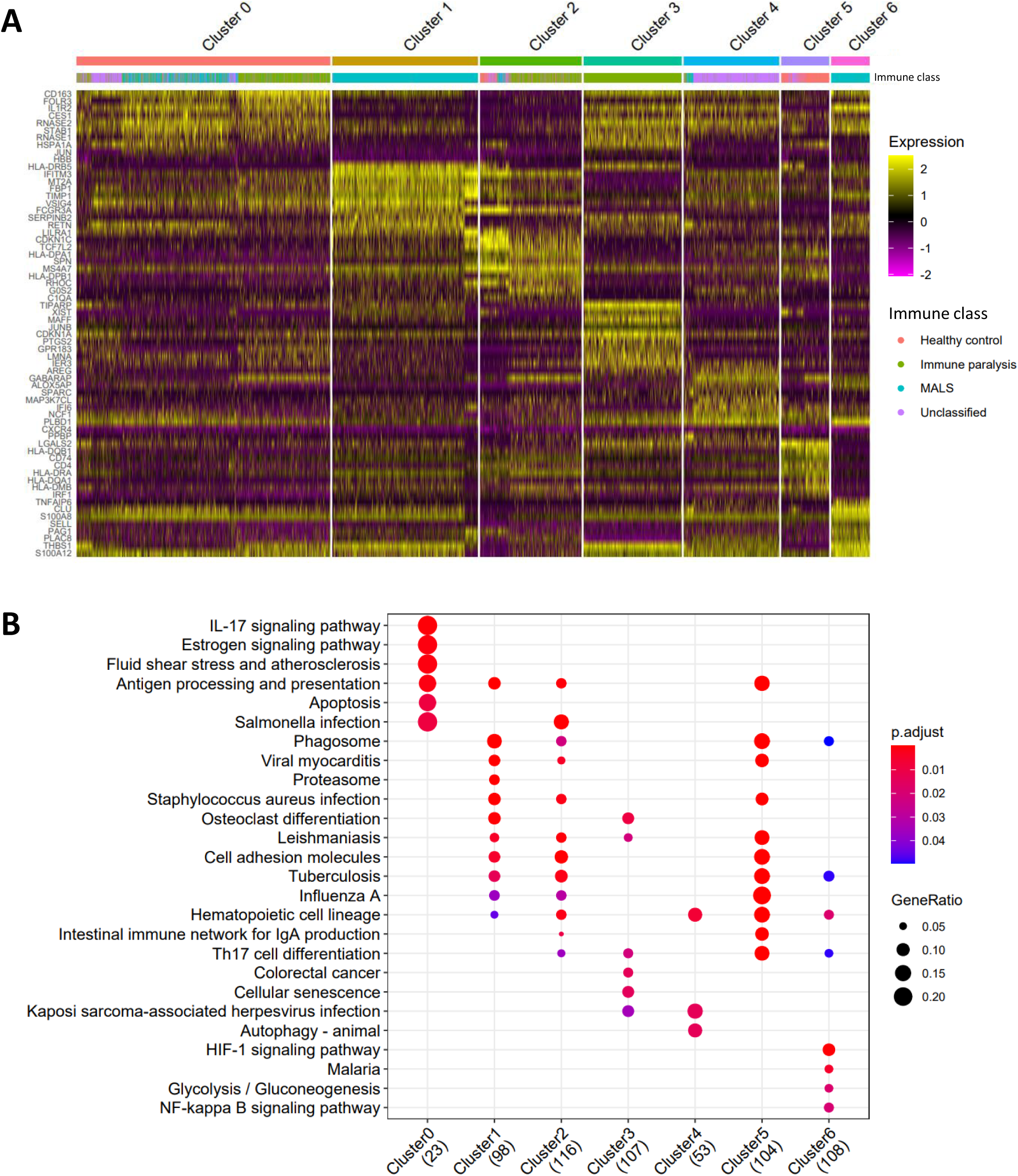
Functional analysis of monocyte subsets. (**A**) Heatmap of the top 10 marker genes of each monocyte cluster. (**B**) Enrichment analysis based on monocyte cluster marker genes using the KEGG database.

### Identification of monocyte cluster signatures in an *in-vitro* endotoxin tolerance model

In order to uncover the role of the novel monocyte subpopulations identified, we aimed to link our identified monocyte populations to relevant cell signatures from experimental models. To that aim, we performed an additional study in an experimental model of immune paralysis induced by endotoxin tolerance in human primary monocytes (15) (**Figure 6A**). In short, tolerization was induced by exposing monocytes to LPS for 24 hours, after which they were left to rest for 5 days, followed by restimulated with LPS for an additional 4 hours. Transcriptome profiles of monocytes were measured by using single cell SORT-seq technique (16). The differentially expressed genes between tolerant (LPS pre-exposed) and control (unexposed/naive) cells after restimulation with LPS were determined, after which the enrichment of this signature was calculated for each monocyte subset (**Figure 6B**). Interestingly, the *in-vitro* LPS-induced immune tolerance signature was strongly present in the transcriptome of monocytes from cluster 6 (mainly found in MALS patients). This suggests that MALS is also associated with poor immune responsiveness, describing the current thoughts on the concept of immune dysregulation in sepsis, which is characterized by concurrent excessive inflammatory and immune suppressive features (17).

**Figure 6:**
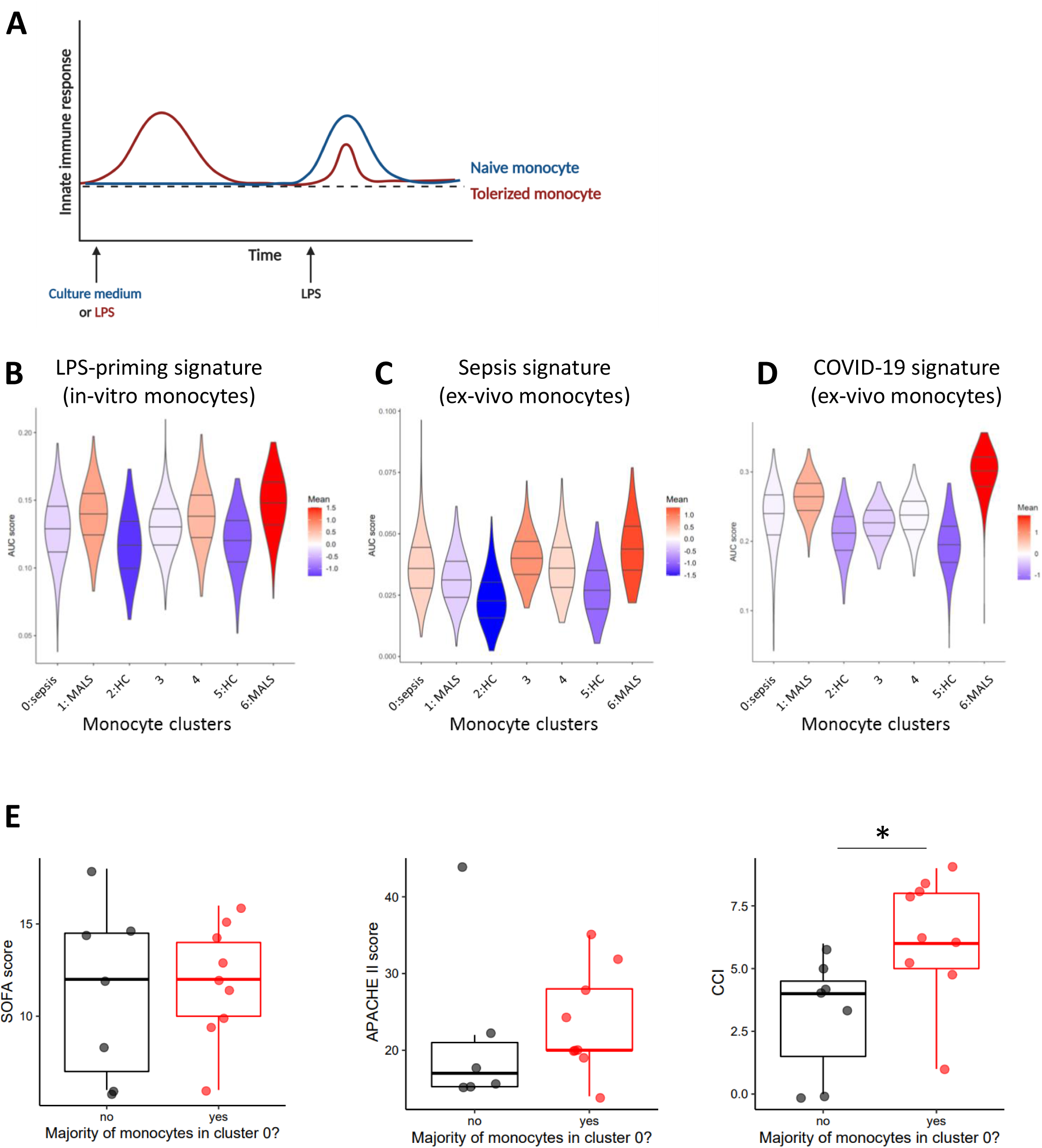
Validation of monocyte subsets and linked to disease. A graphical overview of LPS-induced tolerance in monocytes *in vitro* (**A**). A gene set enrichment analysis to test for the enrichment of gene signatures of an *in-vitro* LPS tolerance model (**B**), bacterial sepsis (**C**) and severe COVID-19 (**D**) in the marker genes of the monocyte subsets identified in this sepsis cohort. The area under the curve (AUC) values are visualized in violin plots. (**E**) Boxplots comparing four disease severity markers (the Sequential Organ Failure Assessment [SOFA] score, the Acute Physiology and Chronic Health Evaluation [APACHE] II score, and the Charlson Comorbidity Index [CCI]) between sepsis patients having the majority of their monocytes as part of cluster 0 versus sepsis patients who do not have the majority of their monocytes in cluster 0. * p-value<0.05.

### Validation of monocyte cluster signatures in urosepsis and COVID-19 patients

In a next set of analyses, we assessed the signature of a previously reported sepsis-specific monocyte cluster described in sepsis patients due to urinary tract infections (18). This signature was especially present in cluster 3 and 6 (**Figure 6C**), which were enriched in immune paralysis (cluster 3) or MALS (cluster 6). This demonstrates once more the immune heterogeneity of sepsis, and that the anatomical/etiologic stratification of sepsis patients is not enough to identify immune endotypes and guide immunotherapy. Interestingly, both the *in-vitro* LPS immune tolerance signature as well as the bacterial sepsis signature were absent in cluster 2 and 5, which are the clusters that are mainly present in healthy controls.

Next, we analyzed a monocyte signature specific for severe COVID-19 disease, based on genes that are higher expressed in monocytes from severe COVID-19 patients compared to patients with mild disease (19). Interestingly, transcriptional signatures of monocytes from cluster 1 and 6 overlapped with the signature reported in severe COVID-19 disease (**Figure 6D**), which are both MALS-specific monocyte clusters, indicating an overlap in monocyte responses between severe COVID-19 and MALS. Indeed, the hyperinflammatory character of COVID-19 is now well established (20–22), and modulatory immunotherapy with anti-IL-1 or anti-IL-6 treatment has been proven successful (23–26).

Finally, we aimed to better understand the clinical relevance of monocyte subcluster 0, which is present in al immune endotypes of sepsis patients. As shown in **Figure 4E**, there is a clear dichotomy within the sepsis population, where patients either have the majority of their monocytes belonging to cluster 0 or not. We compared these two sets of patients, with either a large (> 70%) or small (< 15%) proportion of their monocytes in cluster 0, with regard to three clinical scores that reflect severity of disease, comorbid conditions or indicate poor prognosis: the Sequential Organ Failure Assessment (SOFA) score, the Acute Physiology and Chronic Health Evaluation (APACHE) II score, and the Charlson Comorbidity Index (CCI; **Figure 6E**). For APACHE II, there is a clear trend visible, where the cluster 0 monocyte population was linked to a higher score. For CCI, this was statistically significant (Wilcoxon rank-sum test, p-value<0.05, adjusted p-value=0.07). These data suggest that the sepsis-specific monocyte cluster 0 probably mirrors disease severity.

## Discussion

Treatment of sepsis, and especially the use of immunotherapy, remains challenging due to the heterogeneity of this patient population, both clinically and immunologically. Both deleterious hyperinflammation (defined as MALS) and immune paralysis have been described in patients with sepsis, sometimes even at the same time or in different phases of disease in the same patient. Although these two immunological endotypes are defined by a small number of biomarkers (e.g. ferritin or cytokine concentrations for MALS, HLA-DR expression on monocytes for immune paralysis), without a comprehensive understanding of the biological mechanisms behind them it is difficult to design the most appropriate therapeutic strategy. In an attempt to understand the underlying biological signatures associated with the different immune endotypes identified in sepsis patients, that could thereafter help the design of personalized immunotherapy, we analyzed the transcriptome of circulating immune cells at a single-cell level. While observing clear differences in cell proportions between sepsis endotypes, there were also pronounced differences detected in the transcriptional signature of monocytes from sepsis patients compared to controls, as well as from MALS compared to immune paralysis patients. The noticeable heterogeneity in the monocyte population in sepsis patients also led to the identification of MALS-specific monocyte clusters, as well as one more broad sepsis-specific monocyte cluster which was present in all sepsis endotypes and related to disease severity. These findings shed light on the heterogeneous immune landscape underlying sepsis and identify the transcriptional signatures of different immune endotypes that can function as therapeutic targets in the future.

Defects in both innate as well as adaptive immune cells are proposed to drive (mal)adaptive immune responses in sepsis patients (27). Lymphopenia is commonly observed in sepsis, and is clinically associated with increased susceptibility to secondary infections and late mortality (28). In sepsis patients, enhanced apoptosis has been observed in circulating B cells, NK cells, and CD4 and CD8 T cells (29). Sepsis-induced immune cell apoptosis was also observed in post-mortem patient studies and confirms to be an important mechanism for lymphocyte depletion (30). In line with previous findings, our cell proportion analysis revealed a decrease in lymphocyte subsets CD4 and especially CD8 T cells in patients with severe sepsis. Importantly, we show that this decrease in CD4 T cells was even more pronounced in sepsis patients that were classified as immune paralysis. This is an important observation confirming that not solely the innate immune system is compromised (low monocytic HLA-DR expression), but showing that also adaptive immunity is impaired in these patients, reflecting two important aspects in the pathogenesis of sepsis-induced immune paralysis (27).

Comparing transcriptional profiles of sepsis patients with healthy controls mainly revealed pronounced differences in CD4 T cell and monocyte subsets. In general, an increase in cytokine signaling pathways was observed (TNF and IL-17 signaling) in CD8 T cells, monocytes and NK cells, as well as an increase in oxidative phosphorylation in the CD4 T cells and monocytes, indicating an increase in metabolic activity in these cells. The enrichment of cytokine signaling pathways is not surprising and in agreement with previous studies, considering that TNF-α and IL-17 have already been described to play a role in the pathophysiology in sepsis (31, 32). In addition, in the cell-cell interaction network analysis, we observed that monocytes mainly respond to ligands from other monocyte subpopulations, indicating that monocytes are likely driving a positive feedback activation loop in sepsis that could be important for host defense, but may overshoot in MALS if exaggerated.

Several important new insights were obtained comparing the transcriptomic signatures of immune cells isolated from patients with very different immune endotypes of sepsis patients; MALS vs. immune paralysis. The pathogenesis of MAS / MALS is still not completely understood, but it is believed to be due to the overwhelming release of inflammatory mediators (33, 34). In genetically-mediated MAS, a primary defect in lymphocyte cytolytic activity of mainly NK cells and CD8 T cells leads to the inability to lyse infected cells, but also the failure of apoptosis of highly activated T-cells and macrophages, eventually resulting in a long-lasting antigen-driven immune activation and further amplification of the pro-inflammatory response (35). In contrast, our data show that MALS differs transcriptionally from other immune endotypes of sepsis (such as immune paralysis) especially by changes in monocytes, rather than lymphoid cells. This finding that myeloid cell hyperactivation is likely responsible for the exaggerated release of inflammatory mediators in MALS is in line with the observation that an exaggerated biological activity of the IL-1/IL-18 family of cytokines characterizes hyperinflammation in sepsis. In contrast, immune paralysis is characterized by a low expression of HLA-DR on monocytes and defective responsiveness of both myeloid and lymphoid cells to microbial stimulation (36). Our data also show that immune cells from sepsis patients with MALS have an enrichment of upregulated genes linked to apoptosis in NK cells, CD4 T cells and B cells, and phagosome activation in CD4 T cells, CD8 T cells, B cells and monocytes, compared to immune paralysis patients. Lymphocyte apoptosis is believed to be an important component of immune dysregulation in sepsis (37), which combined with our data suggests that apoptosis inhibitors may have a role in supporting immune function of sepsis patients. Importantly however, our data also suggest that apoptosis pathways are upregulated only in a sub-group of patients with sepsis, and immune stratification of sepsis patients to identify MALS should be strongly considered if modulators of apoptosis are considered.

Our data show that the highest number of differentially expressed genes was observed in the monocyte fraction when comparing MALS to immune paralysis patients. Based on previous data (9), high ferritin levels (>4,420 ng/mL) were used to diagnose patients with signs of MALS in our study. Here, we were able to validate this at a gene-transcriptional level reflected by high expression of *FTL*, a subunit of ferritin, in monocytes. In addition, our pathway analysis showed upregulated genes related to specific intracellular infections (e.g. influenza A and salmonella), hinting towards the interferon pathway, as well as an increase in NADH dehydrogenase pathway. Together, these changes may reflect the plastic nature of monocytes/macrophages during immune activation in MA(L)S, and transcriptional reprogramming may be one of the important mechanisms involved in the pathogenesis of this immunological entity.

Considering the heterogeneity observed within the monocyte population, as well as the clear transcriptional differences between MALS and immune paralysis, we aimed to identify potential subsets of monocytes in the sepsis endotypes. We identified a sepsis-specific monocyte cluster (‘cluster 0’), which was present in all sepsis patients and was related to clinical scores reflecting severity of illness. IL-17 signaling pathway was significantly enriched in this monocyte cluster, which could contribute to local inflammation and tissue damage in sepsis (31), providing a possible explanation for its link with disease severity and a potential future target for therapy. Furthermore, we also identified two monocyte clusters (‘cluster 1’ and ‘cluster 6’) that were uniquely enriched in MALS patients, and absent in the monocytes of sepsis patients with other immune endotypes and healthy controls. Interestingly, metabolic dysregulation targeting glucose metabolism and HIF-1α pathways are strongly enriched in monocyte cluster 6, arguing that restoration of the metabolism of glucose in immune cells of MALS patients could represent an important therapeutic avenue. This hypothesis is supported by earlier studies showing that sepsis in both animal models (38) and patients (39, 40) is accompanied by metabolic dysregulation. The identification of these unique clusters makes increasingly clear that immune heterogeneity is an important characteristic of sepsis on the one hand. This concept was validated here in an independent cohort of patients with urosepsis, showing a mixture of MALS and immune paralysis transcriptional programs. This demonstrates the importance of biomarker-driven application of immunotherapeutic approaches, opening new avenues for personalized immunotherapy.

Finally, the identification of a sepsis-specific monocyte cluster that is linked to higher clinical scores such as APACHE II, SOFA and CCI, all have been liked to increased mortality in sepsis patients (41–43). We observed that monocyte cluster 0 was associated with higher CCI scores, and showed a clear trend with higher APACHE II score, independent of the underlying immune endotype. This suggest that genes specific to this cluster could be used to develop a panel to use in clinical practice to identify patients with severe disease and poor prognosis early on. Importantly, this monocyte cluster was identified in a population of severely ill sepsis patients, and therefore validation of this monocyte cluster in an independent population of sepsis patients is of critical importance.

Our study also has limitations, most important being the small sample size. MALS is a relatively rare immune endotype in sepsis, with only about 5% of sepsis patients displaying this immune profile, and 20% of patients with septic shock (13, 44). Although, immune paralysis is observed to be more frequent (about 40%), relatively large numbers of sepsis patients needed to be screened to be able to identify patients with this immune state. Another limitation of this study is its cross-sectional design; a longitudinal assessment using a multi-omics approach will be needed to provide a complete picture of the highly dynamic immune response shaping the disease course of sepsis. Although the findings were validated in independent cohorts of urosepsis and COVID-19 patients, as well as in a model of experimental LPS-induced tolerance, more studies are needed in order to validate the data of the present study.

In conclusion, we report heterogeneity of monocyte populations in critically ill sepsis patients with different immunological endotypes ranging from MALS to immune paralysis. We identified a sepsis-specific monocyte subset that is linked to disease severity, as well as MALS-specific monocyte gene clusters that could represent the basis for personalized therapeutic targeting. Understanding and identifying monocyte signatures underlying immune endotypes offers a promising approach to stratify patients for precision medicine in sepsis in the future.

## Methods

### Study design

This study includes samples of a subset of sepsis patients who participated in a double-blind, double-dummy randomized clinical trial that took place between November 2017 and December 2019 in Greece, called the PROVIDE study (ClinicalTrials.gov identifier NCT 03332225, EudraCT number 2017-002171-26). The PROVIDE study consisted of one screening/observational stage and one intervention (clinical trial) stage (13). Samples for this study were collected during the screening stage.

### Patient population and inclusion

#### Subjects

The sepsis patients participating in this study were recruited to one of the 14 study sites in Greece, meeting inclusion and exclusion criteria as described before (13). All patients (newly admitted or already hospitalized) included were diagnosed with sepsis or septic shock, according to the Sepsis-3 definition (2), due to community-acquired pneumonia, healthcare-associated pneumonia, ventilator-associated pneumonia, acute cholangitis, or primary bloodstream infection. These infections were defined according to international used criteria (45) and were applied to all patients, as previously described (13). In parallel, control samples were collected from middle-aged healthy individuals without known comorbidities or chronic diseases, and no use of medication.

#### Immunological screening procedures

Patients diagnosed with sepsis due to lung, primary bacteremia, or acute cholangitis were subject to additional screening methods for their immune state. Blood samples were obtained through venipuncture; within the first 24 hours after diagnosis (day 1) and this was repeated after 24 hours (day 2). Samples were collected in EDTA-tubes and transferred via courier to the central laboratory. Ferritin was measured daily in serum by an enzyme immunosorbent assay (ORGENTEC Diagnostika GmbH, Mainz, Germany; lower detection limit 75 ng/mL). Using whole blood from the EDTA vial, white blood cells were incubated for 15 minutes in the dark with the following monoclonal antibodies: Quantibrite HLA-DR/anti-monocyte PerCP-Cy5 (Becton Dickinson, Cockeysville Md). After incubation, cells were analyzed by flow cytometer FC500 versus cells stained with anti-CD45 PC5 and anti-idiotypic IgG1. HLA-DR molecules on CD14/CD45 monocytes were expressed as a number of molecules per cell.

#### Subgroup definitions

According to the immune classification that was proposed in this study (13), MALS was identified in patients with ferritin concentration >4,420 ng/mL, which was measured on the day on which samples could be obtained (day 1 or day 2), regardless of flow cytometry results. Immune paralysis was defined as HLA-DR/CD14/CD45 expression <5,000 Mab/cell and ferritin concentration was ≤4,420 ng/mL. Patients that could not be classified as MALS or immune paralysis were considered having an intermediate immune status (ferritin ≤4,420 ng/mL and HLA-DR ≥5,000 antibodies/monocyte), so-called “unclassified”.

#### Patient characteristics

Baseline characteristics, clinical data (including source of infection, comorbidities and calculated CCI), and outcome were collected for each patient from medical files. The Acute Physiology and Chronic Health Evaluation (APACHE II) and the Sequential Organ Failure Assessment (SOFA) score were calculated on day 1.

### PBMC isolation and cryopreservation

Peripheral blood mononuclear cells (PBMCs) were isolated from 20 mL EDTA blood samples that were obtained in parallel with immunological screening procedures (day 1 or 2), using optimized density 1.077 g/mL (Lymphosep, Biowest) gradient centrifugation. After three washings in ice-cold PBS (phosphate buffered saline, pH: 7.2), PBMCs were resuspended in RPMI 1640 (Biochrom, Berlin, Germany) enriched with 2 mM of L-glutamine, 100 U/mL penicillin G, 0.1 mg/mL streptomycin. Cells were counted using a Neubauer plate with trypan blue exclusion of dead cells. Next, cells were cryopreserved with a density of 10×10^6^/mL per vial in freeze medium containing 90% FBS and 10% DMSO by using a freezing container (Coolcell) at -80°C overnight. Thereafter, vials were transferred for long-term storage to liquid nitrogen, until further analysis.

### DNA genotyping

DNA was isolated from 0.5 mL blood with the ReliaPrep chemistry (Promega) automated on the Freedom Evo Robot (Tecan). Next, genotyping was performed using the GSA-MDv3 array (Infinium, illumina) with a lab-specific clusterfile. This information on genetic variation was used for demultiplexing of the single-cell RNA-sequencing data.

### Single-cell RNA-sequencing and downstream analyses

#### Sample preparation and sequencing

Frozen PBMCs were thawed according to the instructions given by 10x Genomics in preparation for their Single Cell protocols (Fresh frozen human peripheral blood mononuclear cells for single cell RNA sequencing protocol- Document CG00039). Briefly, frozen PBMCs were recovered by rapidly thawing of cell suspensions at 37°C and immediately sequential dilution (1:1) with pre-warmed RPMI media containing 40% fetal bovine serum (FBS). The dilution step was repeated every minute for a total of 5 times, followed by centrifugation at 300 g for 5 minutes. After centrifugation, cells were resuspended in PBS with 0.04% BSA, washed once, passed through a 40μm mesh cell strainer, followed by determination of cell concentrations and viability using an automated Cell counter (TC20 automated Cell counter, Bio-rad). Next, samples were prepped for scRNA-seq according to the 10X Genomics Chromium™ Single Cell 3′ v2 RNA sequencing specification. Cells from 3-5 samples were pooled for one reaction at a final stock within the recommended range (700-1200 cells/µL) and loaded at a volume with a targeted cell recovery count of 1200 cells for each sample. As a high percentage of non-viable cells may impact the targeted cell recovery in these pools, in case the minimum viability (≥ 70%) was not achieved, non-viable cells were first removed from the individual samples with a removal of dead cell kit according instructions and recommendations of the manufacturer and 10x Genomics (MACS Dead Cell Removal Kit, Miltenyi; removal of dead cells for single cell RNA sequencing-document CG000093) before pooled and loaded. The generated cDNA was used for Illumina next-generation sequencing using a NextSeq500-v2 150 cycle kit with a sequencing depth of 25,000 reads/cell.

#### Reads alignment and demultiplexing

The data were aligned to the GRCh38 reference genome using CellRanger (10X Genomics) to generate count tables. Then, demultiplexing of the samples within each pool was performed using souporcell, which is a method to cluster cells using the genetic variants detected within the scRNA-seq reads (46). Doublets detected by souporcell were excluded from further analysis.

#### Quality control, unsupervised clustering and cell annotation

After demultiplexing, samples were further analyzed using Seurat v3 (47) in R version 4.0.3. Cells were excluded if number of unique genes expressed was < 200 or > 4000 and if the percentage of mitochondrial reads was > 25%. Log-normalization was applied before downstream analysis and the count data was scaled regressing for the percentage of mitochondrial reads. Mitochondrial and ribosomal reads were also excluded from downstream analysis.

Subsequently, principal component analysis was performed using the top 2000 variable genes as input. Then, the first 20 dimensions were used as input for Uniform Manifold Approximation and Projection (UMAP) to cluster cells in an unsupervised manner. Clusters were identified using a shared nearest neighbor (SNN) modularity approach with a resolution of 0.4. These clusters were then annotated to their respective cell types based on the expression of canonical markers.

#### Cell proportion, differential expression and pathway enrichment analysis

Cell proportion differences were calculated using Dirichlet-multinomial regression. Then, per cell type, differentially expressed genes were identified using a Wilcoxon rank-sum test, while no filtering was performed based on the fold difference. These differentially expressed genes were then used for KEGG enrichment analysis separately for up and downregulated genes using the R package clusterProfiler (48). Throughout the study, p-value adjustment was performed using FDR, except for the differential expression analyses, where a Bonferroni correction was applied based on the recommendations from the Seurat R package (47). A p-value<0.05 was considered statistically significant and all tests performed were based on two-sided hypothesis testing.

#### Cell-cell interaction network

A ligand-to-target interaction network was generated based on genes differentially expressed in sepsis compared to healthy controls. The NicheNet algorithm was used with monocytes as the receiver cell type and all the other main cell types as sender cells (49). The ligand-receptor interactions were visualized using the circlize package in R (50).

#### Identification of regulatory transcription factors

To identify the transcription factors regulating the expression profiles of monocytes from the three immunological endotypes in sepsis, we followed the SCENIC workflow (51). The RcisTarget and GENIE3 R packages were used to identify regulons and to reconstruct gene regulatory networks (52). The hg38 (refseq_r80) motifs version 9 database was used to annotate to transcription factors. The genes that were differentially expressed in monocytes between the three immunological endotypes (MALS, unclassified and immune paralysis) were used as target genes.

#### Signature genes of monocyte clusters and gene set enrichment analysis

Due to the heterogeneity in the monocyte population in this study, we selected only the monocytes and performed UMAP on the first 20 dimensions with a resolution of 0.2 to identify clusters within the monocyte population. The marker genes for each cluster were identified using Wilcoxon rank-sum tests, while being expressed in at least 25% of the cells of a cluster with a log fold difference threshold of 0.25. The top 10 marker genes for each monocyte cluster were subsequently visualized in a heatmap.

A gene set enrichment analysis was performed using the R package AUCell (51) to test the enrichment of signatures of an *in-vitro* LPS tolerance model, bacterial sepsis (18) and severe COVID-19 (19) in the marker genes of the monocyte subsets identified in this sepsis cohort. The area under the curve (AUC) values were subsequently visualized in violin plots.

### *In-vitro* LPS tolerance model

#### Sample preparation and stimulation

Isolation of monocytes and induction of immune tolerance by LPS was performed as earlier described (53). Briefly, PBMCs from three donors were isolated by density centrifugation of Ficoll-Paque (GE healthcare, UK), followed by monocyte enrichment by Percoll gradient. Cells were resuspended in RPMI 1640 culture medium (Roswell Park Memorial Institute medium; Invitrogen, CA, USA) supplemented with 2 mM Glutamax (GIBCO), 1 mM pyruvate (GIBCO), and 50 μg/mL gentamicin. After one hour of adherence, the non-adherent cells were washed away with PBS, and the monocytes were incubated with culture medium only (negative control) or 10 ng/mL LPS for 24 hours at 37°C. Then, cells were washed with PBS and incubated for 5 days in culture medium supplemented with 10% human pooled serum, after which the monocytes were restimulated with 10 ng/mL LPS at day 6 for 4 hours.

#### Cell sorting and single-cell RNA-sequencing

Monocytes were sorted from all samples based on forward and side scatter, and well-based scRNA-seq using SORT-seq platform was performed, which is based on the CEL-Seq2 protocol (16, 54). Viable single cells were sorted into 192 or 384-well plates and transcriptome of each well containing a single cell was sequenced.

#### Reads alignment and quality control

First, pair-end reads were processed with the SORT-seq pipeline. Fastq reads were aligned to the human transcriptome (GRCh38) with the Burrows-Wheeler Alignment tool (55). The output of this pipeline is a digital gene expression (DGE) matrix for each sample, which records the number of unique molecular identifiers (UMIs) for each gene that are associated with each cell barcode. Genes detected in less than three cells were removed from the DGE matrix. Cells were also filtered if the number of detected genes was less than 100 or more than 7000, to avoid empty wells or doublets, and if more than 25% of the gene counts were from mitochondrial genes, to remove dying cells.

#### Unsupervised clustering, cell annotation and differential expression analysis

Seurat v3.1 integration workflow with SCTransform normalization method was used to cluster cells into distinct cell subsets (56). In order to keep the biological differences for downstream analyses, batch correction was only used in the cell clustering related steps. For the other analyses, we used standard LogNormalization methods. Then, differential expressed genes were identified using FindMarkers/FindAllMarkers functions in Seurat v3.1 with the Wilcoxon rank-sum test. For each comparison, genes expressed in at least 10% of the cells per group and Bonferroni corrected P-values < 0.05 were regarded as significantly differentially expressed genes. The gene signature of LPS tolerance was defined as genes detected by differential expression analysis between LPS-restimulated macrophages of tolerized macrophages (primed with LPS) versus LPS-restimulated macrophages of RPMI-controls (primed by RPMI) in the *in-vitro* experiments.

### Statistics

The statistical analyses of the baseline and clinical characteristics were performed using Kruskal-Wallis tests, assuming non-Gaussian distribution of variables, for continuous variables, and Fisher-Freeman-Halton tests for nominal variables. A p-value < 0.05 was considered statistically significant and Bonferroni correction was applied to correct for multiple testing. The analysis of the single-cell RNA-sequencing is described in detail (see the two previous sections). All statistical analyses were performed using IBM statistics, SPSS version 25 or R/Bioconductor (R version 4.0.3; https://www.R-project.org/).

### Study approval

From all study subjects, informed consent was obtained from the participant or from their legal representatives prior to participation (approval by the National Ethics Committee of Greece 78/17).

### Code availability

Code generated to process the data are freely available on Github (https://github.com/CiiM-Bioinformatics-group/Sepsis).

### Data availability

All data will be made available on the European Genome-Phenome Archive (EGA; ega-archive.org). The accession code is being generated and will be added once it becomes available.

### Author contributions

E.J.G.B, Y.L. and M.G.N. designed the study. I.G. designed and performed the experiments, and helped with the interpretation of the data. V.A.C.M.K. performed the scRNA-seq analysis, prepared the figures and helped with the interpretation of the data. A.K., N.A., G.D. helped with the laboratory experiments. W.L. and B.Z. helped with the computational data analysis. C.J.X. and Y.L. supervised the data analysis. C.J.X, E.J.G.B, Y.L. and M.G.N. oversaw the study and helped with the interpretation of the data. I.G. and V.A.C.M.K. wrote the first draft of the manuscript, and all authors provided their input and agreed to the final version of the manuscript.

## Data Availability

All data produced in the present study are available upon reasonable request to the authors.

## Acknowledgements

The authors would like to extend their gratitude to all study participants. Moreover, the authors would like to thank Katrin Rabold, Helga Dijkstra, Heidi Lemmers, and Yunus Kuijpers for technical assistance in performing the experiments and data analysis. This study was supported by a Horizon 2020 ImmunoSep grant (#847422). Y.L. was supported by an ERC starting Grant (948207) and a Radboud University Medical Centre Hypatia Grant (2018). C.J.X. was supported by Helmholtz Initiative and Networking Fund (1800167). M.G.N. was supported by an ERC Advanced Grant (833247) and a Spinoza Grant of the Netherlands Organization for Scientific Research. E.J.G.B. has received honoraria from AbbVie USA, Abbott CH, Brahms GmbH, InflaRx GmbH, MSD Greece, Sobi and XBiotech Inc.; independent educational grants from Abbott, bioMérieux Inc, Johnson & Johnson, InflaRx GmbH, Sobi; and funding from the Horizon2020 Marie-Curie Project European Sepsis Academy (granted to the National and Kapodistrian University of Athens), and the Horizon 2020 European Grant ImmunoSep and RISCinCOVID (granted to the Hellenic Institute for the Study of Sepsis).

## Figure legends

## Notes

### Competing Interest Statement

E.J. Giamarellos-Bourboulis has received honoraria from AbbVie USA, Abbott CH, Brahms GmbH, InflaRx GmbH, MSD Greece, Sobi and XBiotech Inc.; independent educational grants from Abbott, bioMerieux Inc, Johnson & Johnson, InflaRx GmbH, Sobi; and funding from the Horizon2020 Marie-Curie Project European Sepsis Academy (granted to the National and Kapodistrian University of Athens), and the Horizon 2020 European Grant ImmunoSep and RISCinCOVID (granted to the Hellenic Institute for the Study of Sepsis).
M.G. Netea is a scientific founder of Trained Therapeutix Discovery, and has received research grants from Trained Therapeutix Discovery, GSK, ViiV Healthcare and Ono Pharma. The other authors have declared that no conflict of interest exists.

### Clinical Trial

NCT 03332225

### Author Declarations

From all study subjects, informed consent was obtained from the participant or from their legal representatives prior to participation (ethical approval by the National Ethics Committee of Greece 78/17).

