## Supplemental Figures for "Single-cell transcriptomics identifies different immune signatures between macrophage activation-like syndrome and immune paralysis in sepsis"

### Supplemental figure 1

A

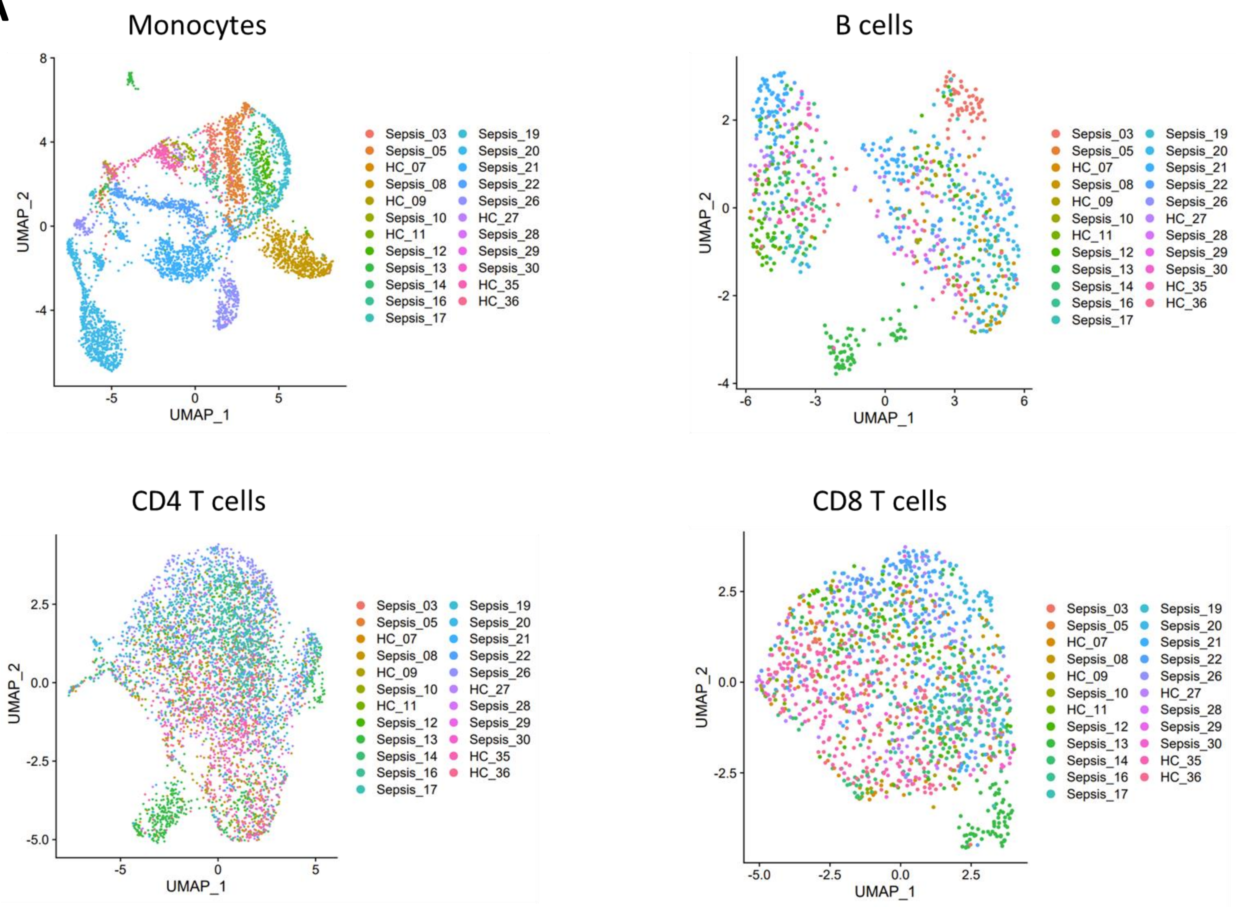


B

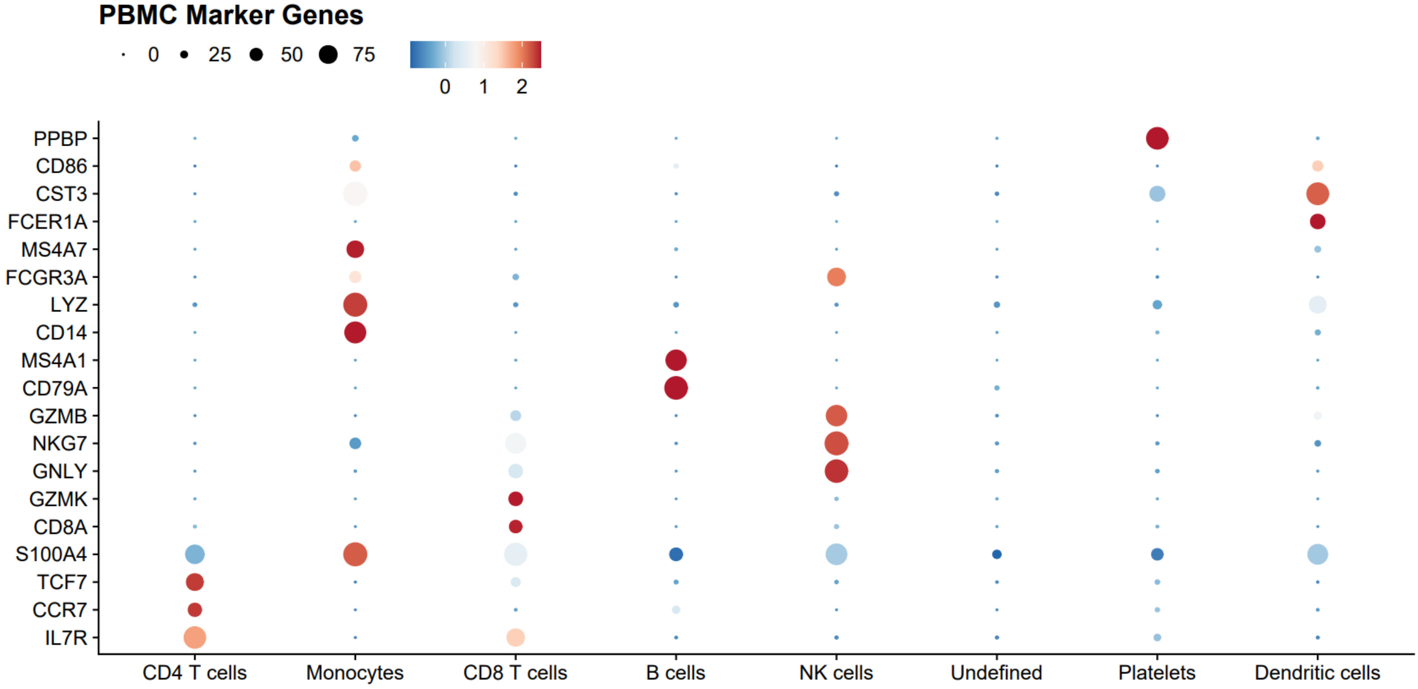

#### Supplemental figure 2: Healthy controls vs. sepsis patients

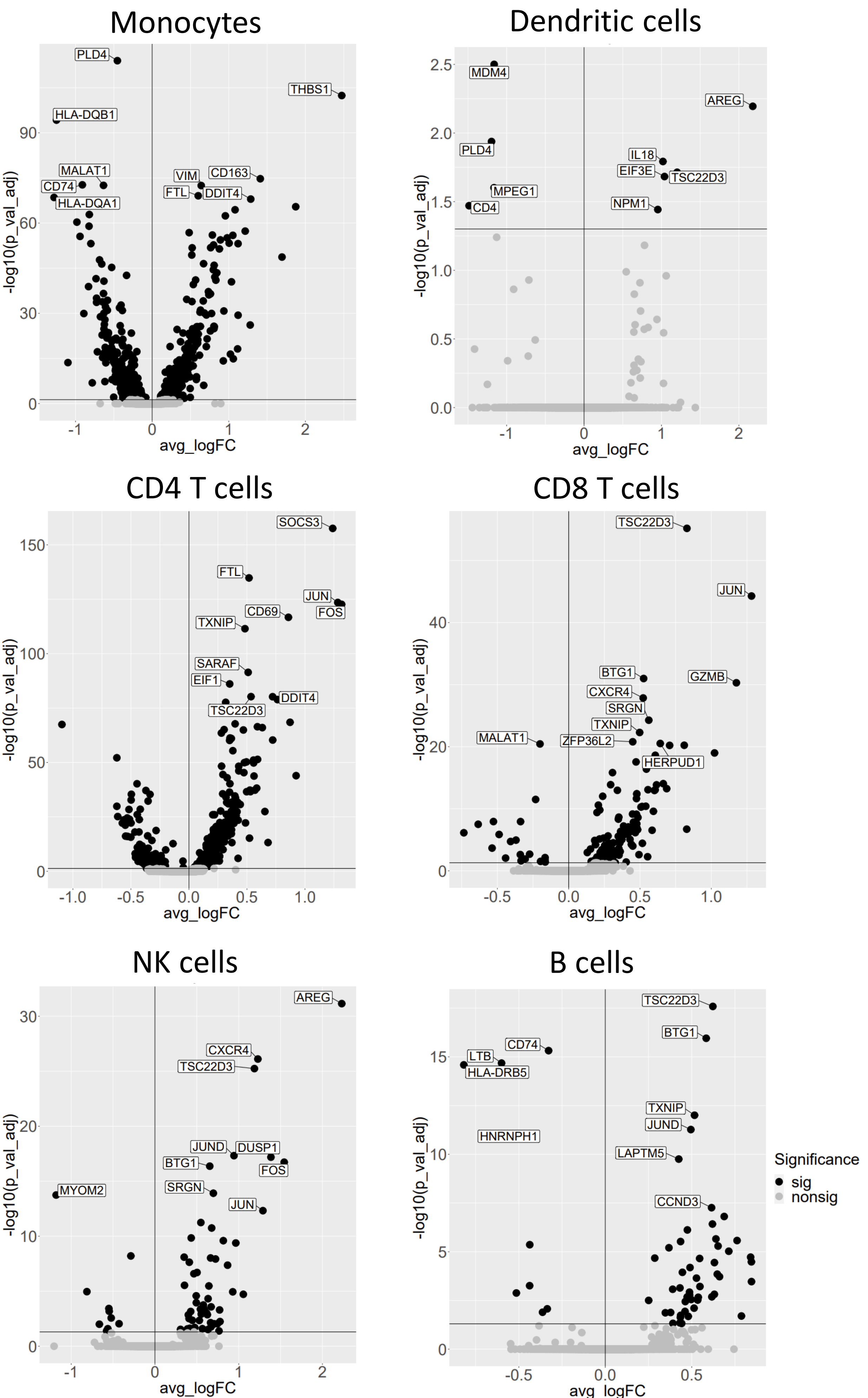

### Supplemental figure 3: Healthy controls vs. sepsis patients (corrected for age and sex)

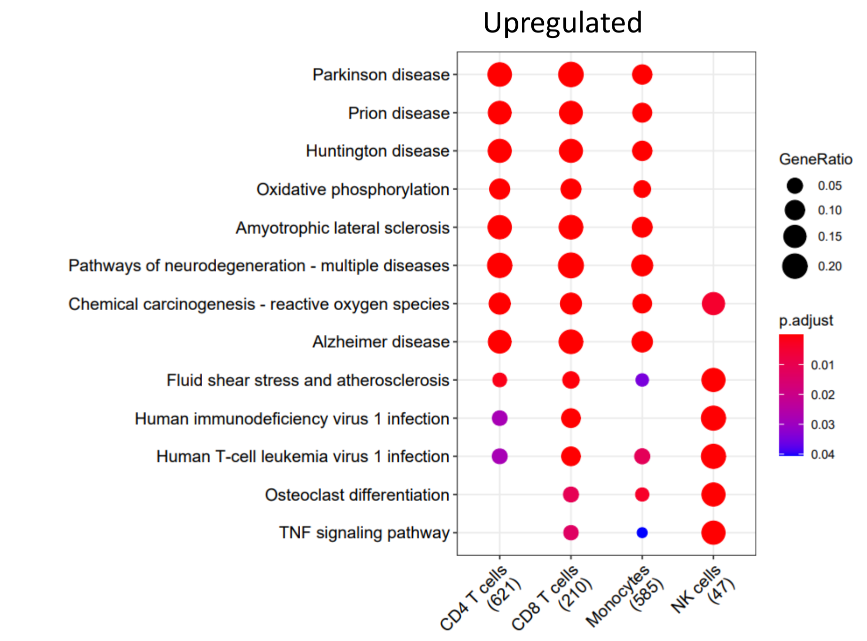


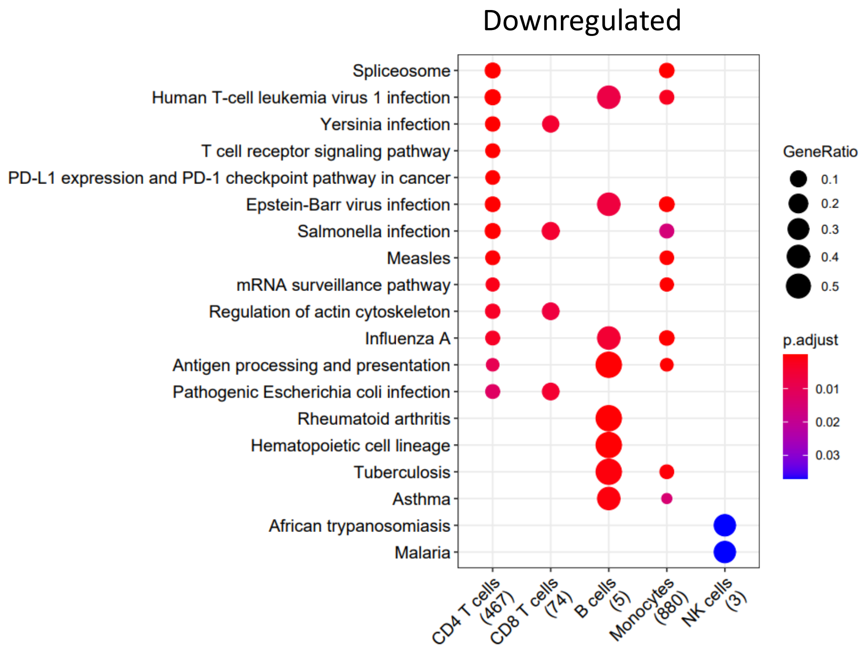

### Supplemental figure 4: Healthy controls vs. sepsis patients (GO terms)

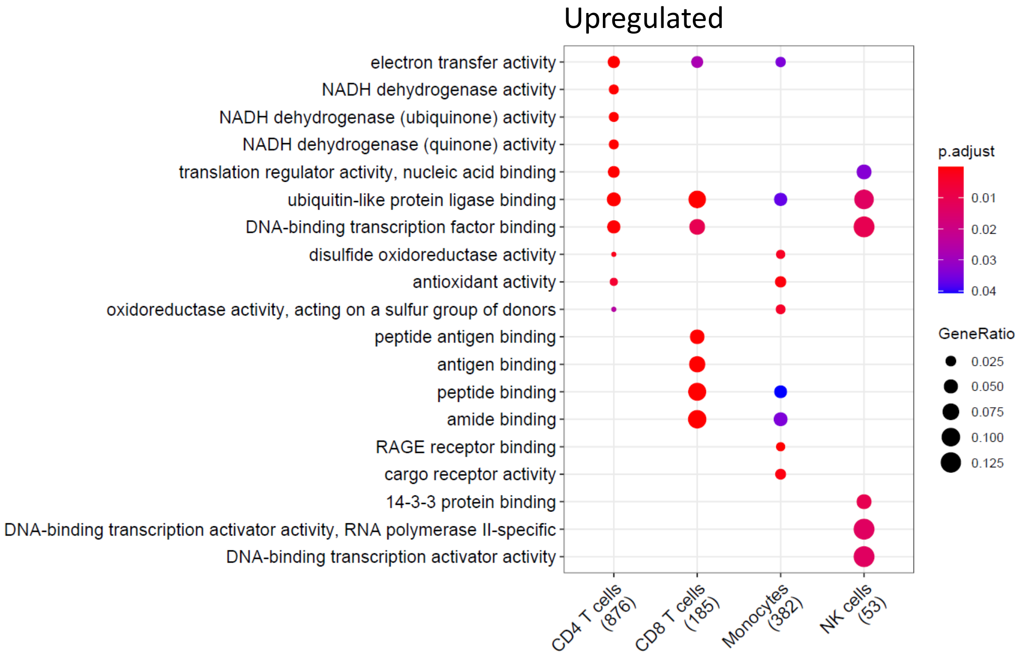


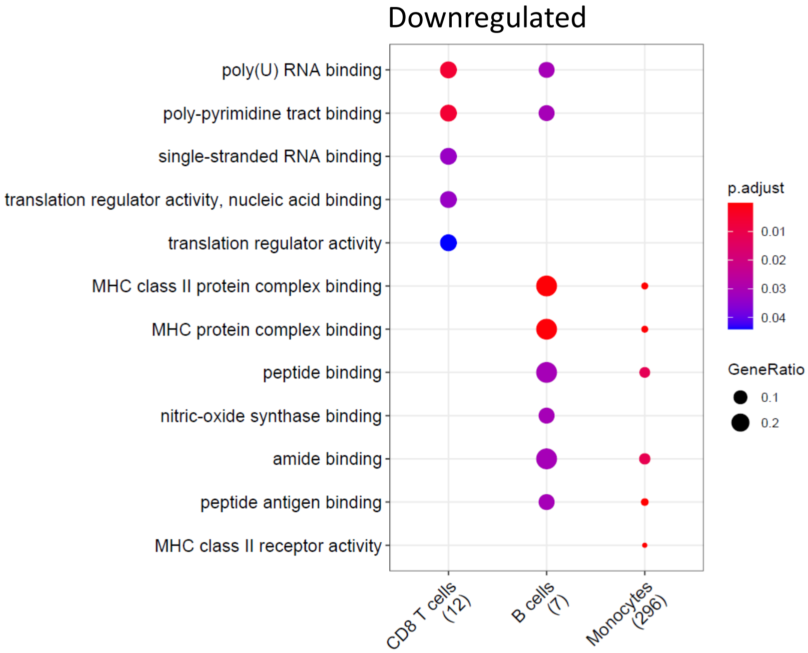

#### Supplemental figure 5: MALS vs. immune paralysis

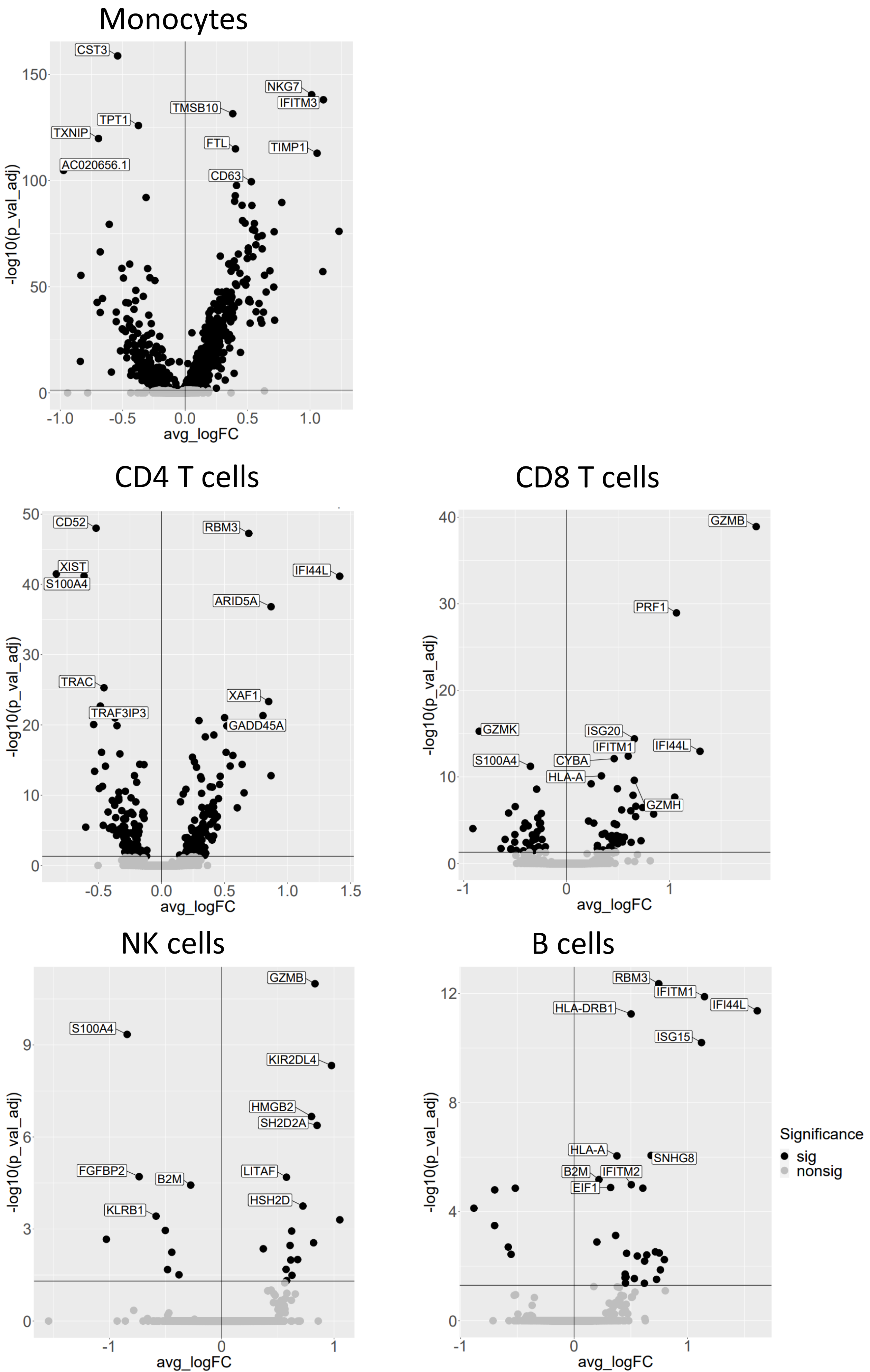

### Supplemental figure 6: MALS vs. immune paralysis (GO terms)

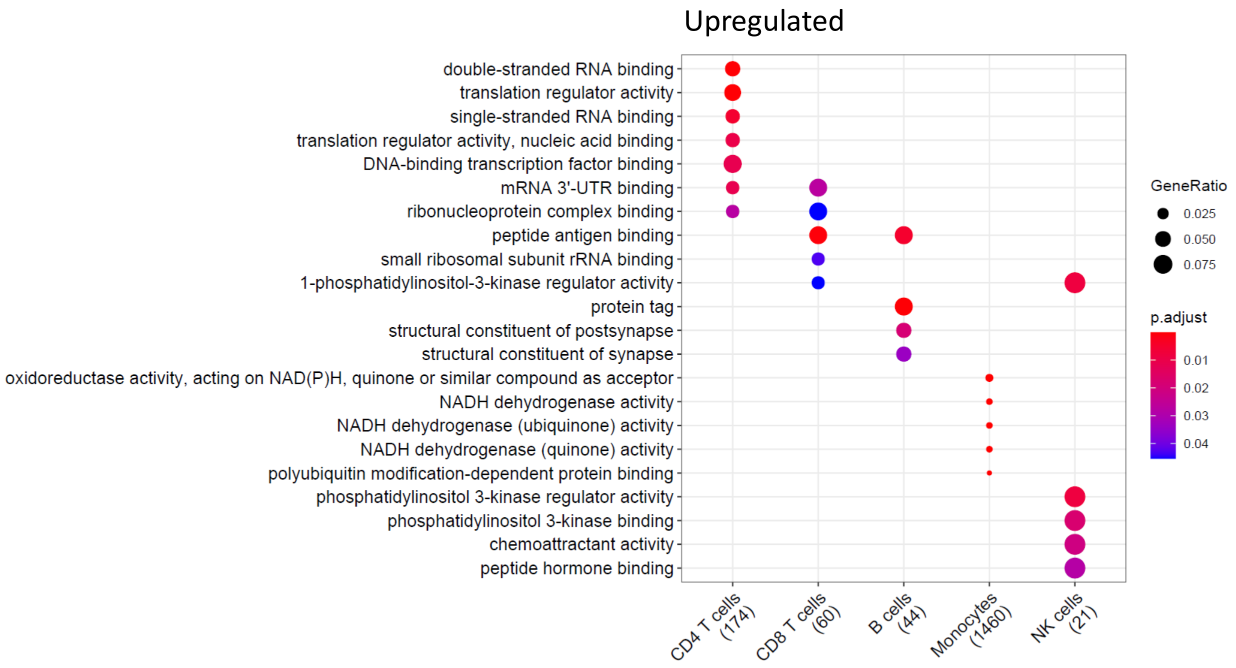


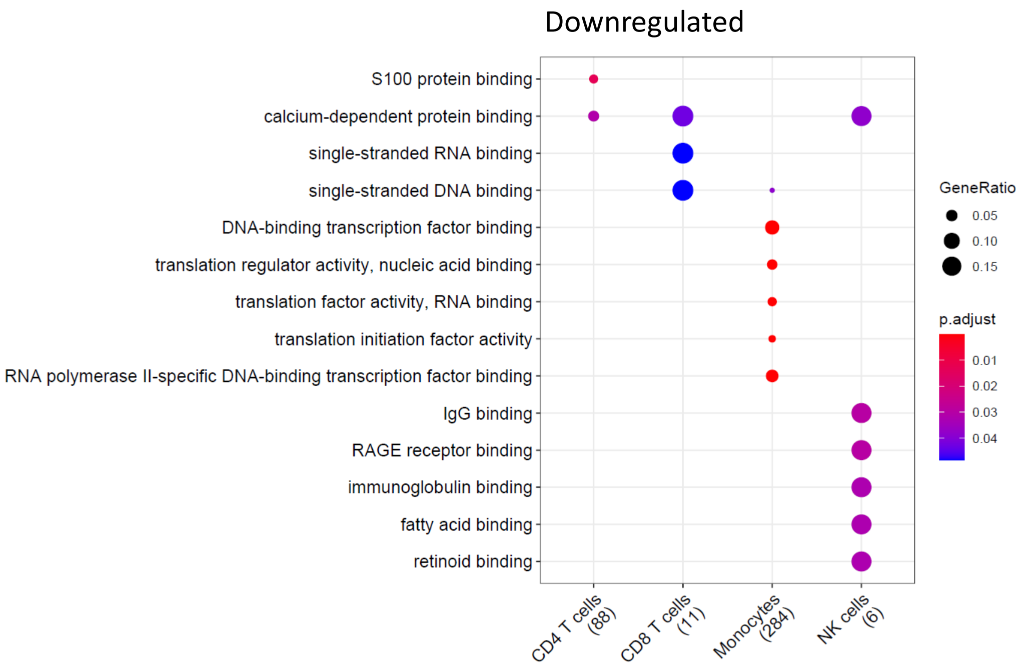

### Supplemental figure 7: Immune paralysis vs. unclassified (GO terms)

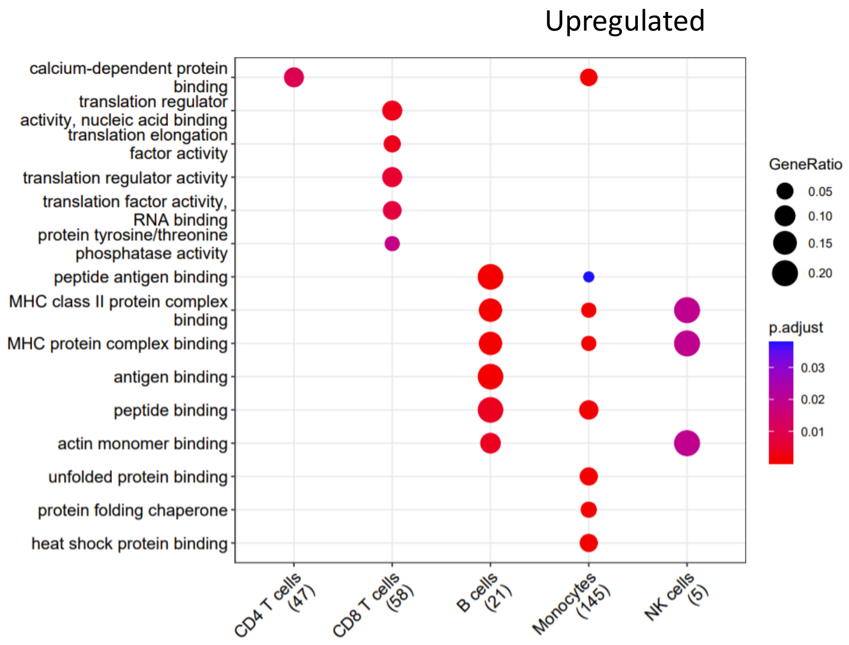


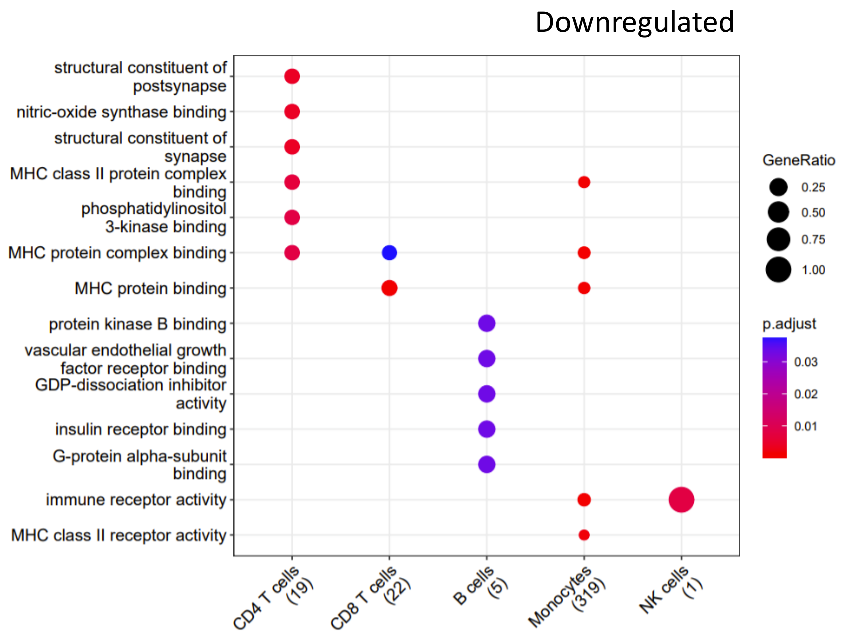

### Supplemental figure 8: Monocyte clustering

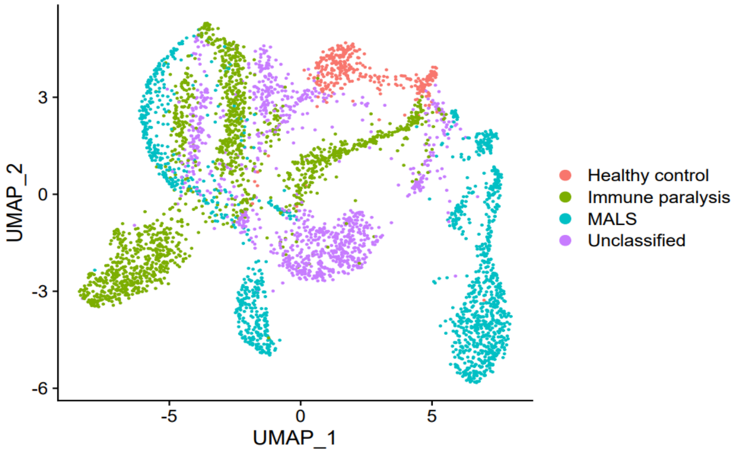


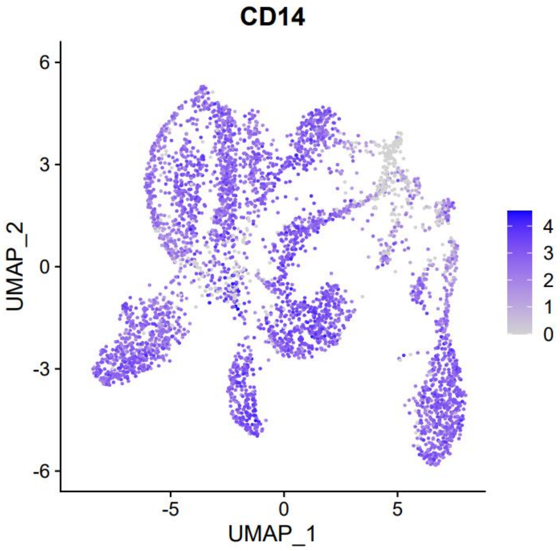


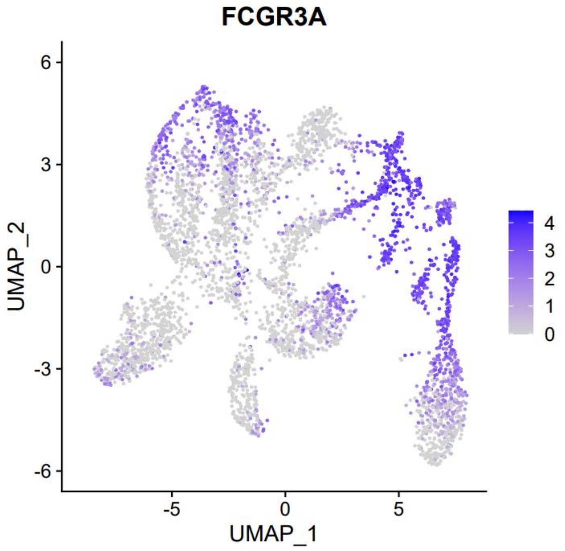

### Supplemental figure 9: Heatmap of monocyte clusters

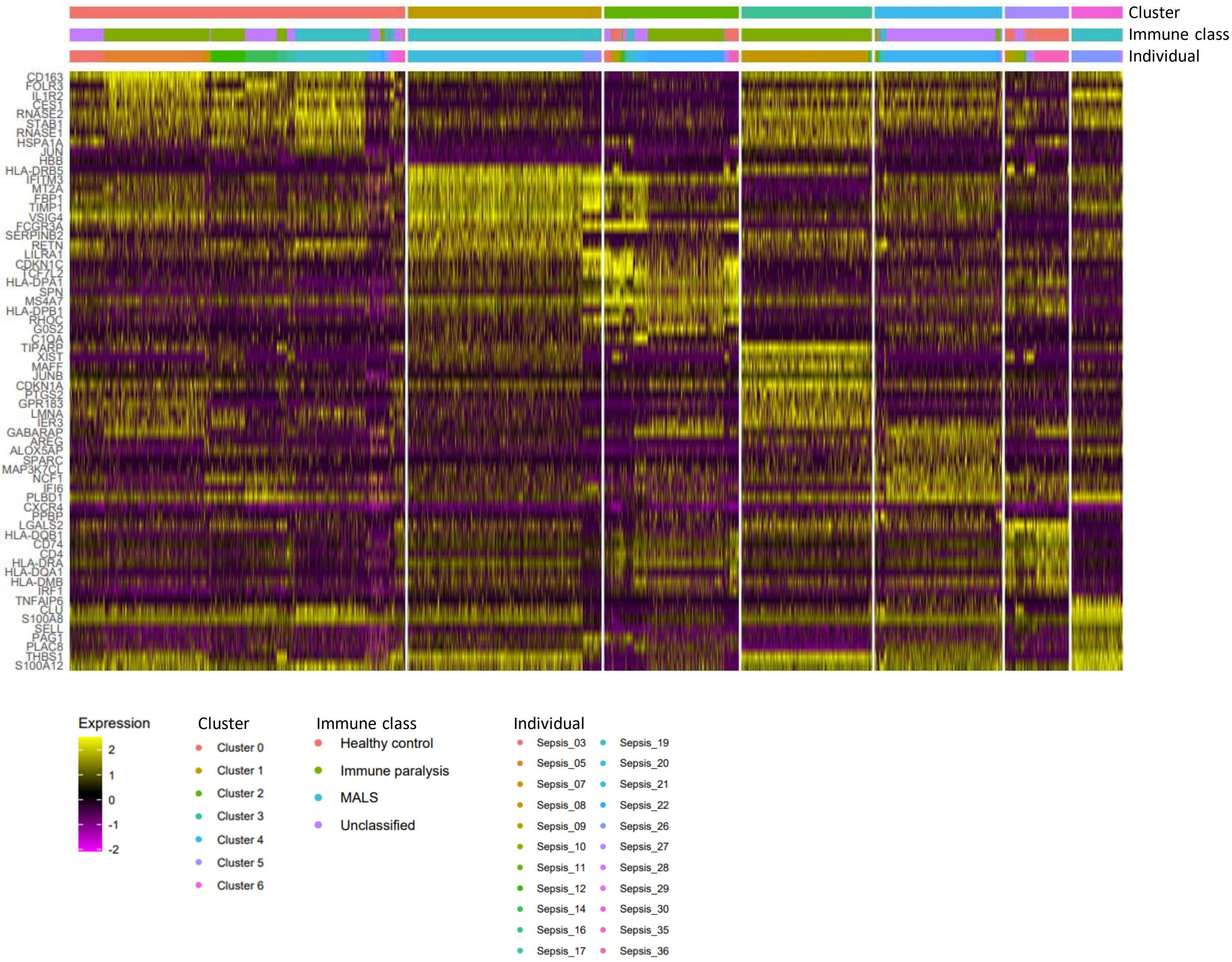
